# Combined Metabolic Activators accelerates recovery in mild-to-moderate COVID-19

**DOI:** 10.1101/2020.10.02.20202614

**Authors:** Ozlem Altay, Muhammad Arif, Xiangyu Li, Hong Yang, Mehtap Aydın, Gizem Alkurt, Woonghee Kim, Dogukan Akyol, Cheng Zhang, Gizem Dinler-Doganay, Hasan Turkez, Saeed Shoaie, Jens Nielsen, Jan Borén, Oktay Olmuscelik, Levent Doganay, Mathias Uhlén, Adil Mardinoglu

**Affiliations:** Science for Life Laboratory, KTH - Royal Institute of Technology, Stockholm, Sweden; Department of Clinical Microbiology, Dr Sami Ulus Training and Research Hospital, University of Health Sciences, Ankara, Turkey; Department of Infectious Diseases, Umraniye Training and Research Hospital, University of Health Sciences, Istanbul, Turkey; Genomic Laboratory (GLAB), Umraniye Training and Research Hospital, University of Health Sciences, Istanbul, Turkey; School of Pharmaceutical Sciences & Key Laboratory of Advanced Drug Preparation Technologies, Ministry of Education, Zhengzhou University, Zhengzhou, Henan, PR China; Department of Molecular Biology and Genetics, Istanbul Technical University, Istanbul, Turkey; Department of Medical Biology, Faculty of Medicine, Atatürk University, Erzurum, Turkey; Centre for Host-Microbiome Interactions, Faculty of Dentistry, Oral & Craniofacial Sciences, King’s College London, London, United Kingdom; Department of Biology and Biological Engineering, Chalmers University of Technology, Gothenburg, Sweden; Department of Molecular and Clinical Medicine, University of Gothenburg and Sahlgrenska University Hospital Gothenburg, Sweden; Department of Internal Medicine, Istanbul Medipol University, Istanbul, Turkey; Department of Gastroenterology, Umraniye Training and Research Hospital, University of Health Sciences, Istanbul, Turkey

**Keywords:** COVID-19, Combined Metabolic Activators, proteomics, metabolomics, omics data

## Abstract

There is a need to treat COVID-19 patients suffering from respiratory problems, resulting in decreased oxygen levels and thus leading to mitochondrial dysfunction and metabolic abnormalities. Here, we investigated if a high oral dose of a mixture of Combined Metabolic Activators (CMA) can restore metabolic function and thus aid the recovery of COVID-19 patients. We conducted a placebo-controlled, open-label phase 2 study and a double-blinded phase 3 clinical trials to investigate the time of symptom-free recovery on ambulatory patients using a mixture of CMA consisting of NAD+ and glutathione precursors. The results of both studies showed that the time to complete recovery was significantly shorter in the CMA group (6.6 vs 9.3 days) in phase 2 and (5.7 vs 9.2 days) in phase 3 trials. A comprehensive analysis of the blood metabolome and proteome showed that the plasma levels of proteins and metabolites associated with inflammation and antioxidant metabolism are significantly improved in patients treated with the metabolic activators as compared to placebo. The results show that treating patients infected with COVID-19 with a high dose of CMAs leads to a more rapid symptom-free recovery, suggesting a role for such a therapeutic regime in the treatment of infections leading to respiratory problems.

## 1 INTRODUCTION

Many patients infected with COVID-19 suffers from respiratory problems, metabolic dysfunction and exacerbated inflammation that triggers organ failure(*1*). This has led to more than 2 million COVID-19 related deaths globally (*2*) due to severe outcomes (*3-7*). Although new treatments regimes have been introduced (*8, 9*), including Remdesivir and Favipiravir (FP) (*10, 11*), there is a need to complement existing therapeutic interventions with new approaches, in particular those targeting mitochondrial dysfunction and metabolic abnormalities. An interesting approach is to promote metabolic function by metabolic activators involved in mitochondrial function.

Metabolic abnormalities and a hyperinflammatory response are associated with the deficiencies in nicotinamide adenine dinucleotide (NAD^+^) and glutathione (GSH) metabolism (*12, 13*). The cellular depletion of NAD^+^ and GSH may be primary factors related to the COVID-19 and the risk for mortality. Given the lack of targeted treatments for emerging viruses, the antiviral properties of repurposed drugs and the use of dietary supplements have gained considerable attention. N-acetyl-L-cysteine (NAC), nicotinamide riboside (NR), and L-carnitine have been tested in human trials of viral diseases including COVID-19, and serine has been evaluated in the treatment of immune system-related disorders (Table S1). Serine is an essential metabolite for modulation of adaptive immunity by supporting the effector T-cell responses (*14*). Many studies have shown that these metabolic activators are beneficial in treating lung diseases and viral infectious diseases (*15-24*). Their pharmacological properties, side effects, and dosing regimens are known, which are advantageous for rapid testing in clinical trials and COVID-19 treatment protocols (*24, 25*).

Previously, we found that combined metabolic activators (CMA) consisting of L-serine (serine), NAC, NR, and L-carnitine tartrate (LCAT, the salt form of L-carnitine) is a potential treatment for nonalcoholic fatty liver disease based on integrative analysis of multi-omics data derived from different metabolic conditions (*24, 26-28*). We subsequently demonstrated the safety of CMA in animals and humans (*29*). That study also revealed that CMA significantly improves plasma levels of metabolites associated with antioxidant metabolism and inflammatory proteins.

We therefore hypothesized that administration of CMA would improve metabolic conditions associated with COVID-19, increase the level of NAD^+^ and GSH, activate the mitochondrial metabolism in liver and other tissues, and potentially accelerate the recovery of COVID-19 patients or reduce its severity. To test this hypothesis, we conducted a randomized, controlled, open-label, placebo-controlled phase-2 study to evaluate the efficacy, tolerability, and safety of CMA in ambulatory COVID-19 patients. Based on the favorable findings from the phase-2 study, we conducted a randomized, controlled, double-blinded phase-3 study to evaluate the efficacy, tolerability, and safety of CMA in ambulatory COVID-19 patients. Here, we report the results from the phase-2 and phase-3 clinical trial, including the effect on the time to recovery which was the primary endpoint for both studies. The results are complimented with an extensive analysis of a large number of biochemical parameters, based on both protein and metabolite analysis.

## 2 RESULTS

### 2.1 CMA Accelerates Recovery of COVID-19 Patients in Open-Label Phase-2 Clinical Trial

In the phase-2 study, we recruited 100 adults with a confirmed positive PCR test for COVID-19. Five patients dropped out for personal reasons, and two were hospitalized before administrating the CMA. Of the 93 remaining patients, all of whom completed the study, 71 were randomly assigned to the CMA group and 22 to the placebo group (Figure 1A, Dataset S1A); all patients also received standard of care therapy (for Turkey) with hydroxychloroquine (HQ) for five days. On Days 0 and 14, we assessed the clinical variables and analyzed the differences between Day 0 and Day 14 in the CMA and placebo groups (Dataset S2A).

**Figure 1.**
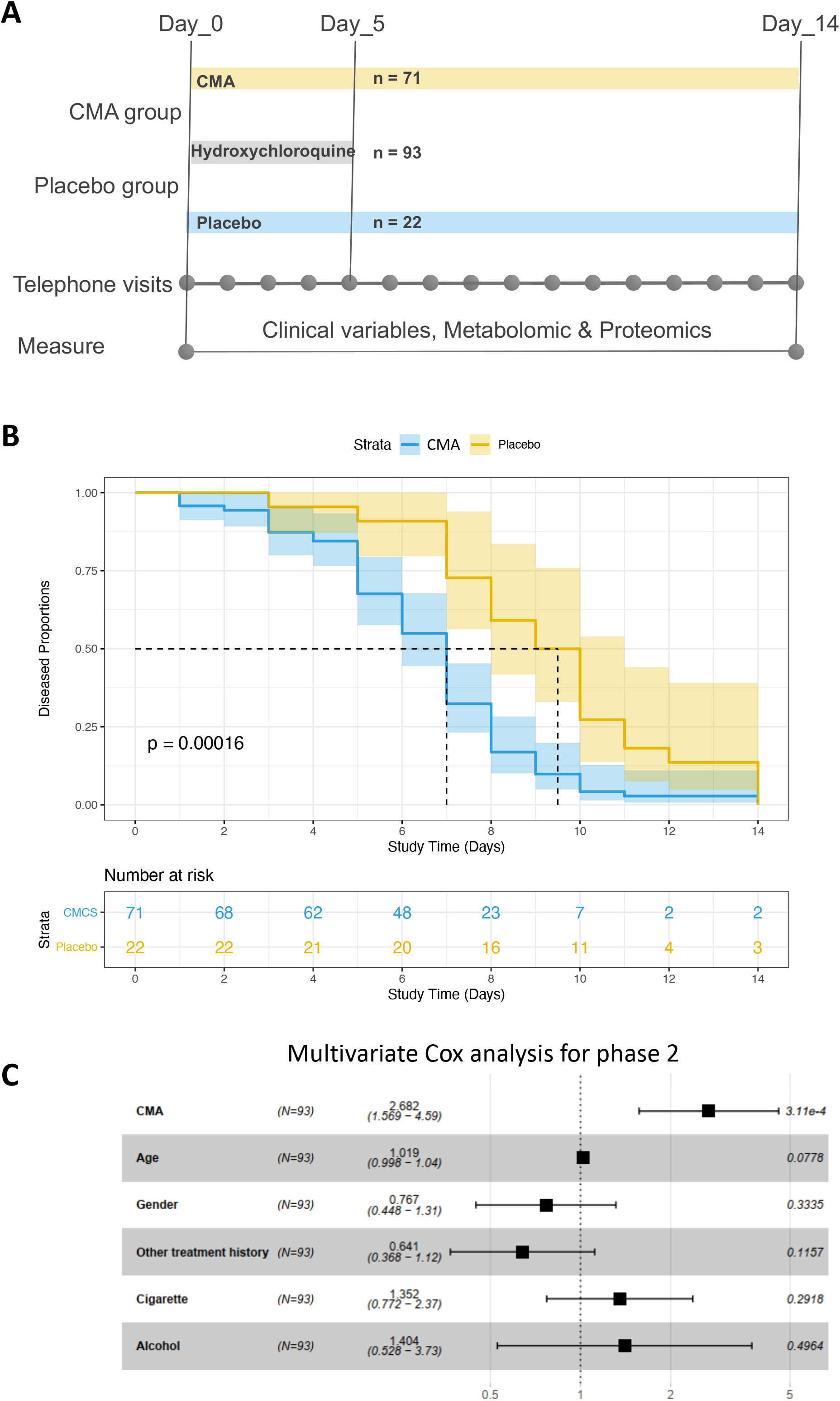
The effect of CMA on the symptoms of COVID-19 patients in phase-2 clinical trial. **A)** The effect of the CMA in COVID-19 patients is tested in a randomized, controlled, open-label, placebo-controlled phase 2 clinical study. Study design for testing the effect of CMA is presented. **B)** Based on Kaplan-Meier analysis, it is shown that CMA accelerate the recovery of the COVID-19 patients. Number of patients reporting COVID-19 symptoms are reported in the Table S2. **C)** Multivariate Cox regression analysis of CMA treatment in phase-2 clinical trial.

The patients’ mean age participated in the phase-2 study was 35.6 years (19–66 years), and 60% were men (Table S2). Patients had a low prevalence of coexisting conditions such as hypertension (2.1%) and type 2 diabetes mellitus (5.3%), and the mean body mass index (BMI) was 24.8 (16.8–37.8). The most common COVID-19 symptoms were tiredness (51.6%), headache (48.3%), cough (31.1%), muscle or joint pain (60.2%), smell or taste disorder (26.8%), sore throat (23.6%), fever (20.4%), breathing issues (6.4%), nausea or vomiting (5.3%), and diarrhea (2.1%). The baseline demographic and clinical characteristics were similar in the CMA and placebo groups (Table S2, Dataset S1A & Dataset S2A). All patients who fully recovered had a negative PCR test for COVID-19 on Day 14. With regards to safety, no severe adverse events occurred and two patients in the CMA group (2.8%) reported adverse events. Both had a mild rash on the upper body and decided to complete the study (Dataset S1A). Adverse effects were uncommon and self-limiting.

In the phase-2 study, we observed that the mean recovery time (the primary outcome variable) was shorter in the CMA group than in the placebo group (6.6 vs 9.3 days, p = 0.0001) (Figure 1B). A univariate Cox regression analysis shows that CMA was significantly associated with recovery time (p < 0.0004, hazard ratio 2.59, 95% confidence interval 1.54–4.38) (Table S3). Multivariate Cox regression analysis analyses also confirmed that CMA independently reduced recovery time after adjustment for age, gender, other treatment histories, smoking, and alcohol consumption (p < 0.0004, hazard ratio 2.68, 95% confidence interval 1.57–4.59) (Figure 1C).

### 2.2 CMA Accelerates Recovery of COVID-19 Patients in Double-Blinded Phase-3 Clinical Trial

In the phase-3 study, we recruited and randomly assigned 309 adult patients with a confirmed positive PCR test for COVID-19. Three patients dropped out for personal reasons, one patient in the CMA group dropped due to mild allergy, and one patient in the placebo group was hospitalized. Of the 304 remaining patients, all of whom completed the study, 229 were in the CMA group and 75 in the placebo group (Figure 2A, Dataset S2B). 62 patients in the placebo group and 187 patients in the CMA group received standard therapy with HQ for five days. Meanwhile, 13 patients in the placebo group and 42 patients in the CMA group received standard therapy with FP for five days. On Days 0 and 14, we assessed the clinical variables and analyzed the differences between Day 0 and Day 14 in the CMA and placebo groups (Dataset S2B).

**Figure 2.**
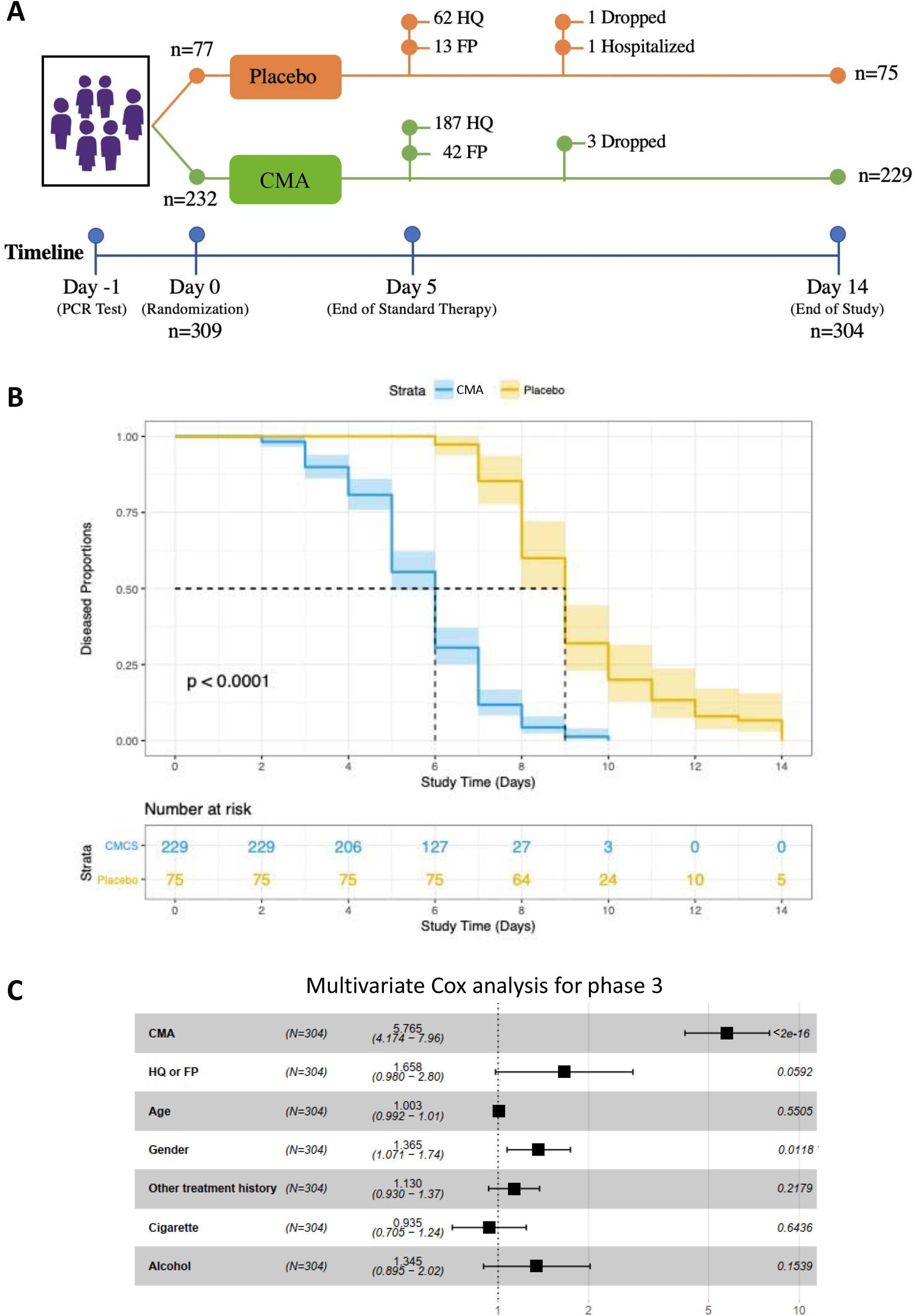
The effect of CMA on the symptoms of COVID-19 patients in phase-3 clinical trial. **A)** The effect of the CMA in COVID-19 patients is tested in a randomized, controlled, double-blinded, placebo-controlled phase 3 clinical study. Study design for testing the effect of CMA is presented. **B)** Based on Kaplan-Meier analysis, it is shown that CMA accelerate the recovery of the COVID-19 patients. Number of patients reporting COVID-19 symptoms are reported in the Table S2. **C)** Multivariate Cox regression analysis of CMA treatment in phase-3 clinical trial.

The mean age of the patients who participated in the phase-3 study was 36.3 years (18–66 years), and 57.6% were men (Table S2). Patients had a low prevalence of coexisting conditions such as hypertension (9.2%) and type 2 diabetes mellitus (6.2%), and the mean BMI was 26.7 (16.8–45.6). The most common symptoms were muscle or joint pain (60.8%), tiredness (50.3%), cough (43.4%), headache (38.1%), fever (28.2%), sore throat (24%), smell or taste disorder (20.7%), breathing issues (9.2%), nausea or vomiting (7.8%), and diarrhea (3.6%). The baseline demographic and clinical characteristics were similar in the CMA and placebo groups (Table S2, Dataset S1B). Two patients in the CMA group (0.6%) reported mild rash and skin redness as adverse events, and both decided to complete the study (Dataset S1B).

In the phase-3 study, we also confirmed that the mean recovery time was shorter in the CMA group than in the placebo group (5.7 vs 9.2 days, p<0.0001) (Figure 2B). A univariate Cox regression analysis shows that CMA was significantly associated with recovery time (p < 2.0e-16, hazard ratio 5.63, 95% confidence interval 4.11-7.71) (Table S4). Multivariate Cox regression analysis also confirmed that CMA independently reduced recovery time after adjustment for HQ/FP treatment, age, gender, other treatment histories, smoking, and alcohol consumption (p < 2.0e-16, hazard ratio 5.77, 95% confidence interval 4.17–7.96) (Figure 2C).

To investigate if the administration of CMA is affected by the standard therapy with HQ and FP, we calculated the mean recovery time in the patient groups treated with HQ and FP separately. We observed that the mean recovery time was again shorter in CMA group than in placebo group (5.76 vs 9.32 days, p<0.0001) in the patients treated with HQ only (Figure S1A) and (5.54 vs 8.77 days, p=0.00034) in the patients treated with FP only (Figure S1B). We also compared the independent effect of the HQ and FP on COVID-19 patients and found that these two drugs had a similar efficacy on the recovery of the patients in all (Figure S2A), placebo (Figure S2B) and CMA (Figure S2C) patient groups. Hence, we observed that CMA administration accelerated the recovery of the COVID-19 patients independent of the standard therapy.

### 2.3 CMA Improves Clinical Health in All COVID-19 Patients

We measured clinical variables in all of the patients recruited in the phase-2 and phase-3 studies and analyzed the differences before and after the administration of CMA in the active and placebo groups (Figure 3A). Analysis of secondary outcome variables showed that serum alanine aminotransferase (ALT) levels were significantly (p = 0.032) lower on Day 14 vs Day 0 only in the CMA group (Figure 3B). We found that the lactate dehydrogenase (LDH) levels (Figure 3C) and creatinine levels (Figure 3D) were significantly lower on Day 14 vs Day 0 only in the CMA group (p = 2.88 E-04 and p = 3.07 E-09), respectively. In contrast, we found that the plasma glucose level was significantly increased on Day 14 vs Day 0 only in the placebo group (Figure 3E). Hence, we observed that the level of ALT, LDH, creatinine and glucose was significantly improved due to the administration of CMA.

**Figure 3.**
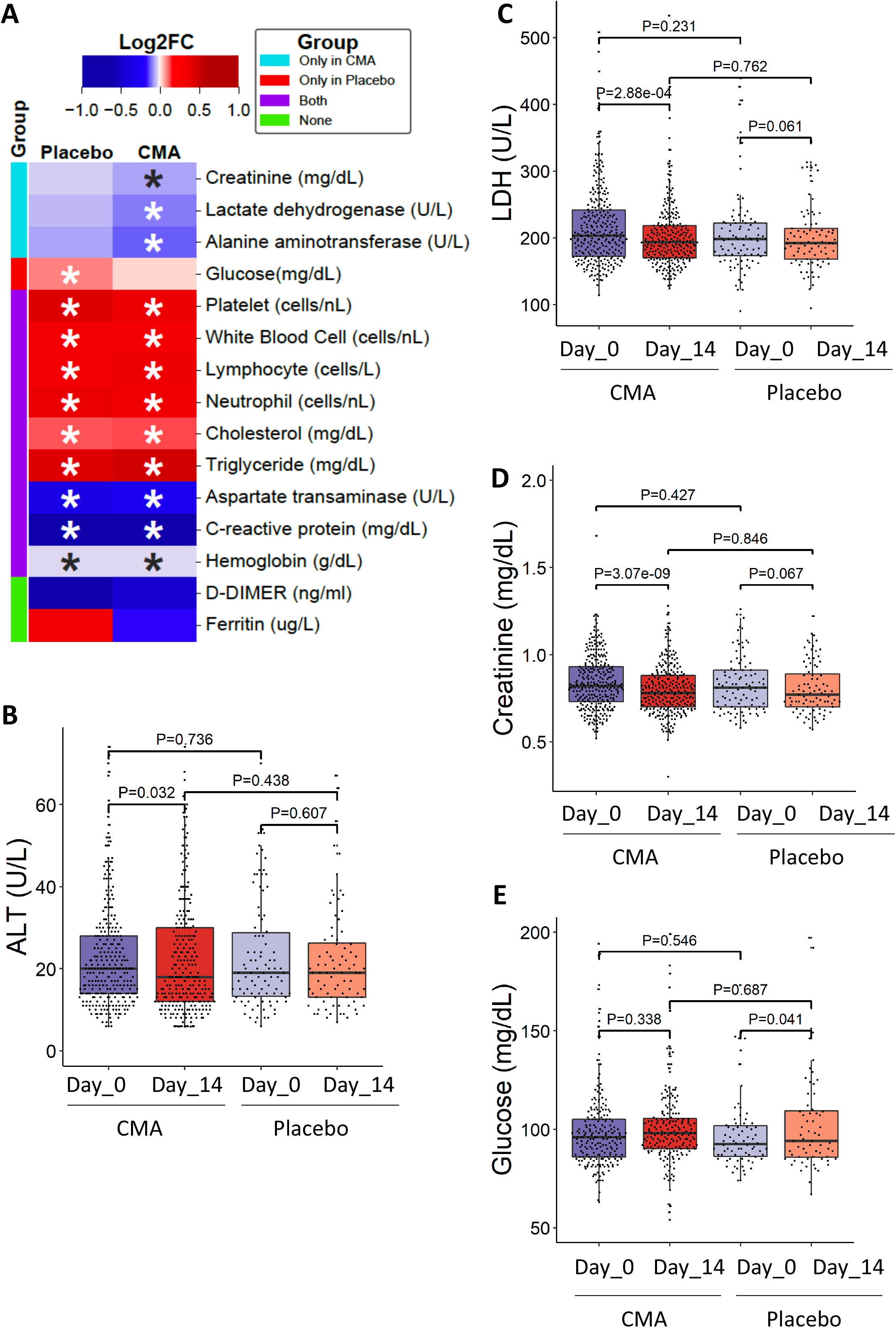
The effect of the CMA on clinical variables. **A)** Heatmap shows log2FC based alterations of the clinical variables, which are compared before and after the administration of CMA in both active and placebo groups. Asterisks indicate statistical significance based on Student’s t test. P value <0.05. The level of clinical variables including ALT **(B)**, LDH **(C)**, creatinine **(D)** and glucose **(E)** is presented in the CMA and placebo groups. These clinical parameters are significantly improved due to the administration of CMA after 14 days. The decrease in the recovery of the patients have been supported by the improvement in clinical variables and the metabolic health status of the patients.

We also measured the level of clinical chemistry parameters and found that their levels are significantly changed in both CMA and placebo groups. We found that the level of aspartate aminotransferase (AST) (Figure S3A) and C-reactive protein (Figure S3B) were significantly lower in both CMA and placebo groups on Day 14 vs Day 0. In contrast, we found that the levels of platelets (Figure S3C), white blood cells (Figure S3D), neutrophils (Figure S3E), lymphocytes (Figure S3F), hemoglobin (Figure S4A), cholesterol (Figure S4B) and triglyceride (Figure S4C) were significantly increased in both CMA and placebo groups on Day 14 vs Day 0. We observed that these differences were due to the course of the disease and not affected by the administration of CMA. D-dimer and Ferritin levels did not significantly differ between both groups on Day 14 vs Day 0 (p > 0.05; Figure 3A, Dataset S2C). Our analysis suggested that administration of CMA improved the clinical parameters in parallel to the decrease in recovery time.

### 2.4 Metabolomic and Inflammatory Cytokine Analysis for Characterization of COVID-19 Patients

We characterized the metabolome and inflammatory cytokine markers of 93 COVID-19 patients involved in the phase-2 study and revealed changes associated with the disease and administration of CMA at the molecular level. We analyzed the plasma level of metabolites and inflammatory protein markers on Day 0 and Day 14 in both groups. Generation of omics data may unveil the underlying molecular mechanisms associated to the differences before and after the disease as well as the differences in the placebo and CMA groups. We generated plasma metabolomics data and analyzed the plasma levels of 1,021 different metabolites (Dataset S3). After filtering out the metabolites with missing values in more than 50% of samples, the plasma metabolomics dataset included 928 metabolites that were subjected to statistical analysis. Significant differences in plasma metabolite levels on Day 14 vs Day 0 in both the CMA and placebo groups (Dataset S4) and between the groups on Day 14 and Day 0 (Dataset S5) are presented. We investigated the associations between the plasma level of all metabolites with the plasma levels of the metabolic activators as well as degradation products of HQ (Dataset S6).

We also measured the plasma levels of 96 inflammatory protein markers using an inflammation panel to quantify the plasma level of target proteins (Dataset S7). After quality control and exclusion of proteins with missing values in more than 50% of samples, 72 proteins were analyzed. Significant differences in plasma protein levels between Day 14 vs Day 0 in both CMA and placebo groups (Dataset S8) and between the CMA and placebo groups on Day 14 and Day 0 (Dataset S9) are presented. We investigated the associations between the plasma level of all inflammation-related proteins with the plasma levels of metabolic activators (Dataset S10). The results showed that the administration of CMA effected a large number of biochemical parameters, based on both metabolomics and proteomics analysis.

### 2.5 CMA Increases the Plasma Levels of Metabolites Associated with Metabolic Activators

We first analyzed the plasma levels of serine, carnitine, NR, and cysteine. CMA increased the plasma levels of each metabolic activator proportionally on Day 14 vs Day 0 in the CMA group (Figure 4A, Dataset S4). Moreover, the plasma levels of NR, nicotinamide, 1-methylnicotinamide and N1-methyl-2-pyridone-5-carboxamide (associated with NR and NAD^+^ metabolism); of serine and N-acetylglycine (associated with serine and glycine metabolism); and of deoxycarnitine, acetylcarnitine, (R)-3-hydroxybutyrylcarnitine, linoleoylcarnitine (C18:2), and dihomo-linoleoylcarnitine (C20:2) (associated with carnitine metabolism) were significantly higher in the CMA group than in the placebo group on Day 14 (Dataset S5).

**Figure 4.**
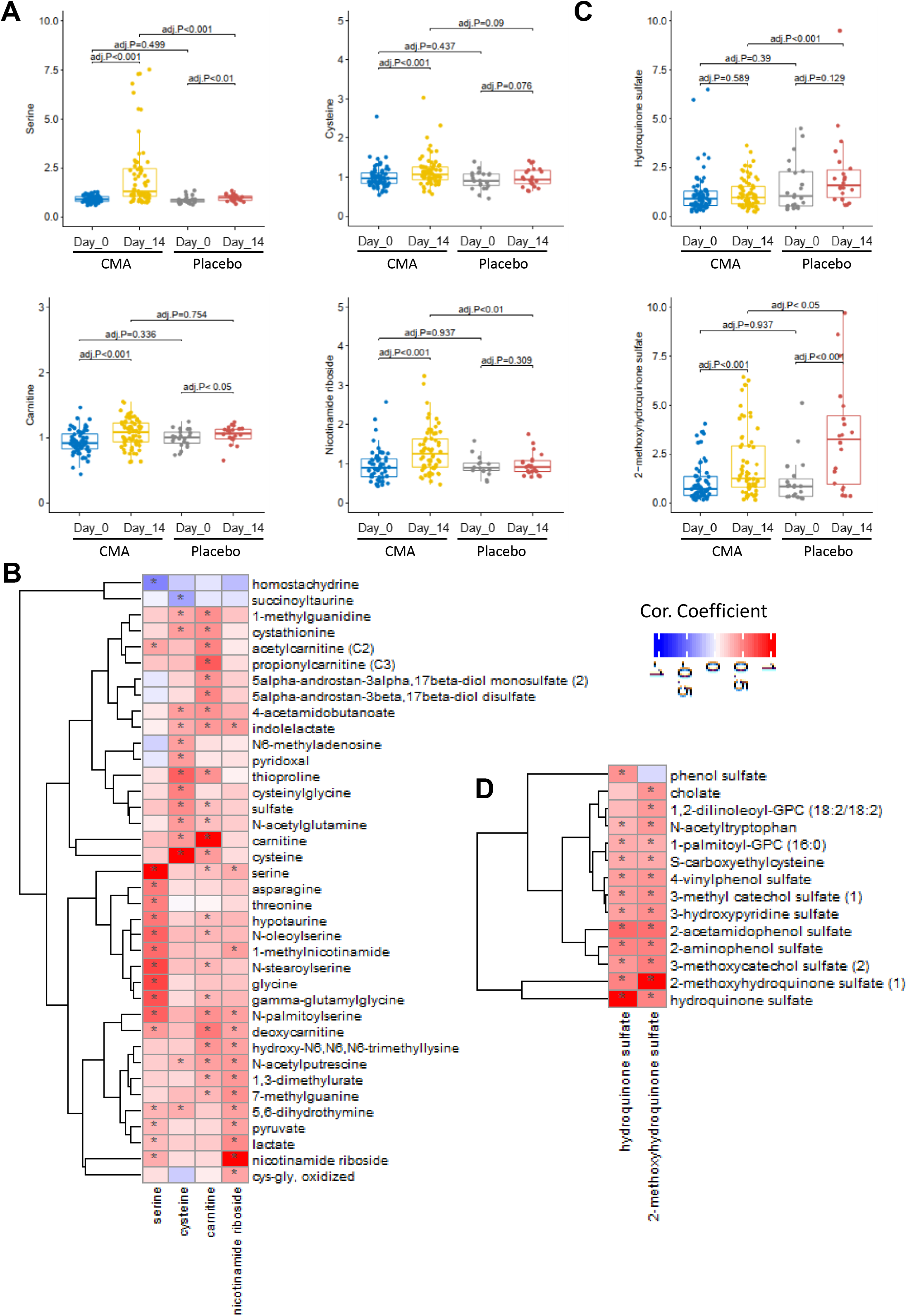
CMA and hydroquinone sulfate alter the global metabolism of the patients. **A)** The plasma level of individual metabolic activators including serine, carnitine, nicotinamide riboside, and cysteine as well as **B)** their association with the 10 most significantly correlated plasma metabolites are presented. **C)** The plasma level of the degradation products of hydroxychloroquine including hydroquinone sulfate and 2-methoxyhydroquinone sulfate (1) as well as **D)** their association with the 10 most significantly correlated plasma metabolites are presented. Asterisks indicate statistical significance based on Spearman correlation analysis. FDR value <0.01.

Next, we investigated the relationship with the plasma level of administrated metabolic activators and other metabolites (Dataset S6). We analyzed the 38 most significantly correlated plasma metabolites with serine, L-carnitine, NR, and cysteine (Figure 4B, Dataset S6) and found three clusters of metabolites which are significantly correlated with serine only, cysteine only and significantly correlated with all serine, L-carnitine, and NR. We observed that cysteine had a different dynamic compared to the other three metabolic activators. On the other hand, homostachydrine involved in xenobiotics metabolism as well as succinoyltaurine involved in methionine, cysteine, SAM, and taurine metabolism were significantly negatively correlated with serine and cysteine, respectively. We also found that homostachydrine, which was significantly positively correlated hydroquinone sulfate (degradation product of hydroxychloroquine), was significantly downregulated on Day 14 vs Day 0 in the CMA group (false-discovery rate [FDR]=1,51E-25, Dataset S4) as well as in the CMA group compared to placebo group on Day 14 (FDR=1,24E-10, Dataset S5).

COVID-19 patients both in the CMA and placebo groups received HQ for 5 days. In order to study the effect of HQ on the global metabolism of the patients, we first analyzed the plasma level of the degradation products of HQ including 2-methoxyhydroquinone sulfate and hydroquinone sulfate. We found that the plasma level of these two metabolites was significantly lower in the CMA vs placebo group on Day 14 and the plasma level of 2-methoxyhydroquinone sulfate was significantly downregulated on Day 14 vs Day 0 in the CMA group (Figure 4C). Hence, we observed that administration of CMA effected the utilization of the HQ in COVID-19 patients. Next, we identified the most significantly correlated 12 plasma metabolites with these 2 metabolites and found that the plasma level of 2-methoxyhydroquinone sulfate and hydroquinone sulfate is associated with the sulfated metabolites, N-acetyltryptophan involved in tryptophan metabolism, S-carboxyethylcysteine involved in cysteine metabolism and cholate involved in primary bile acid metabolism (Figure 4D). Based on this plasma metabolomics analysis, we observed that the metabolic changes reported in the CMA group were not due to the treatment of patients with hydroxychloroquine. On the other hand, we observed that CMA effected the utilization of HQ in COVID-19 patients. Our analysis indicated that 2 weeks administration of CMA increased the plasma level of the individual metabolic activators and improved the plasma level of metabolites associated with the anti-oxidant metabolism.

### 2.6 Cytokine Levels During Recovery of COVID-19 Patients

Mortality in COVID-19 patients has been strongly associated with the presence of the cytokine storm induced by the virus. Increased cytokine levels are also linked with increased viral load and the severe form of the disease (*30*). To study the changes in the cytokine levels, we determined the dynamic range of 72 inflammatory proteins in plasma samples with the Olink multiplex inflammation panel (Dataset S7). The plasma level of 14 proteins were significantly (false-discovery rate: FDR<0.01) changed following the recovery of the patients on Day 14 vs Day 0, in the placebo group (Figure 5A, Dataset S8). The plasma levels of 9 proteins including CSF-1, CX3CL1, CXCL10, IFN-gamma, IL-18R1, MCP-2, OPG, TNF, and TRAIL were significantly (FDR<0.01) decreased, whereas the plasma level of remaining 5 proteins including Flt3L, MCP-4, TNFRSF9, TRANCE and TWEAK was slightly but significantly (log2FC<0.15 & FDR<0.01) increased after 14 days in the placebo group treated only with HQ (Figure 5A, Dataset S8). These results demonstrate that the administration of CMA significantly decreased and increased cytokine levels associated with inflammation during the recovery of the COVID-19 patients.

**Figure 5.**
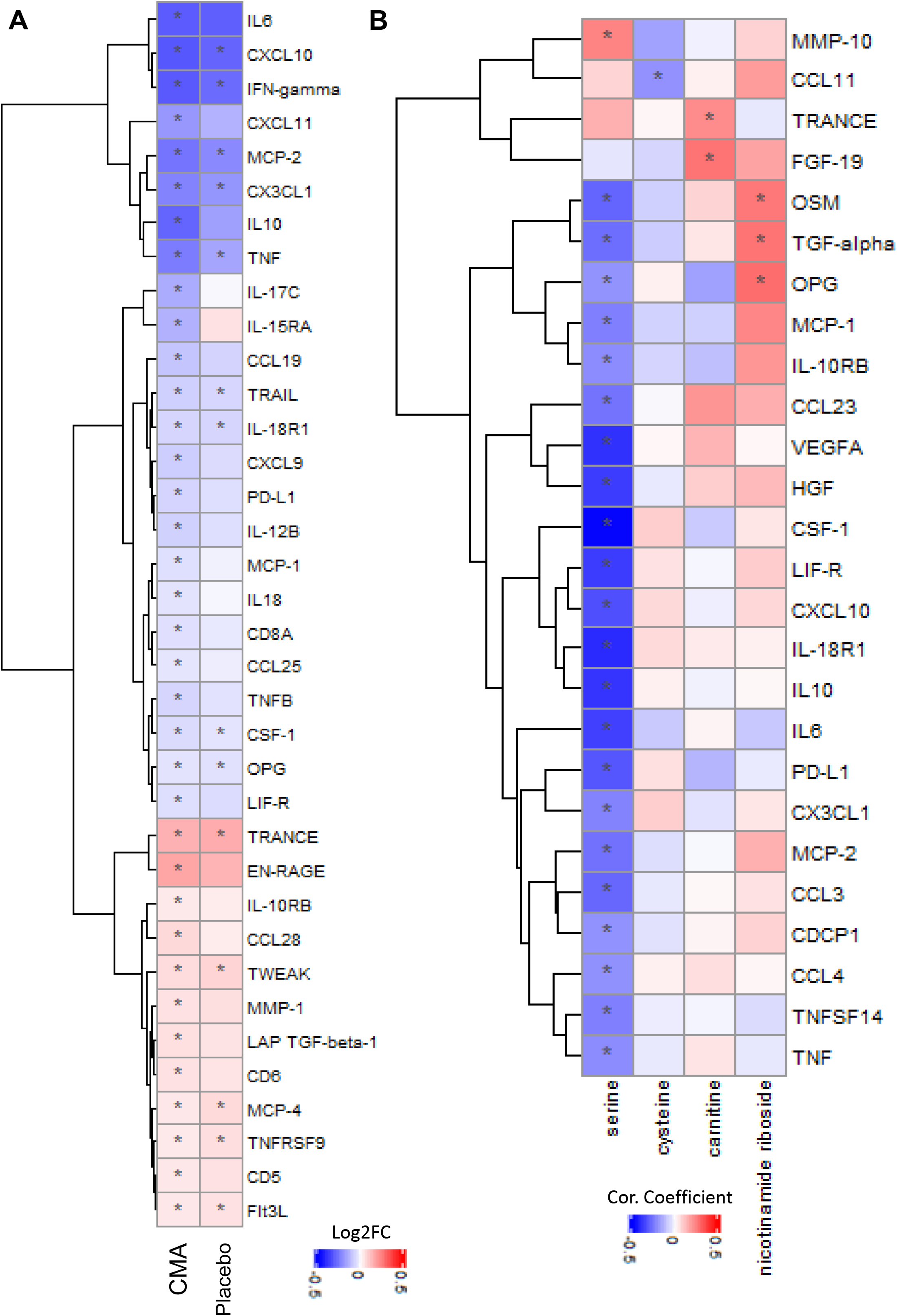
CMA effect the plasma level of inflammation related proteins. Association between inflammation related proteins that are significantly different between the CMA and placebo groups on Days 0 and Day 14. **A)** Heatmap shows log2FC based alterations and clustering between significant proteins on Day 14 vs Day 0 in the CMA and placebo groups. Asterisks indicate statistical significance based on paired Student’s t test. FDR value: < 0.01. **B)** Heatmap shows the correlation and clustering between the plasma level all inflammation related proteins and plasma levels of the individual constituents of CMA (including serine, carnitine, nicotinamide riboside, and cysteine). Asterisks indicate statistical significance based on Spearman correlation analysis. p value: < 0.05. Log2FC: log2(fold change).

### 2.7 Effect of CMA on the Plasma Cytokine Levels

We analyzed the effect of CMA on plasma cytokine levels and found that 36 proteins were significantly (FDR<0.01) different in the CMA group on Day 14 vs Day 0. 12 of these protein markers were significantly increased whereas 24 of these protein markers were significantly decreased on Day 14 vs Day 0. We observed that 14 of the 36 proteins were also significantly (FDR<0.01) differentially expressed in the placebo group (Figure 5, Dataset S8). After filtering out the proteins based on log2FC, we found that the plasma levels of 11 proteins including, IL6, IL10, IFN-gamma, CXCL10, CCL19, CX3CL1, CXCL11, IL-15RA, IL-17C, MCP-2 and TNF were significantly (FDR<0.01) downregulated and the plasma level of TRANCE and EN-RAGE was significantly (FDR<0.01) upregulated in the CMA group (Figure 5 & Dataset S8).

Next, we compared the differences in the decrease of the cytokine levels both in CMA and placebo groups and observed that the magnitude of the decrease in the plasma level of downregulated cytokines was greater in the CMA group compared to placebo group on Day 14 (Figure 6 & Dataset S8). Moreover, we found that the plasma levels of CSF-1, IL-15RA, IL18, MCP-1 and TNF were significantly downregulated in the CMA group compared to placebo group on Day 14, whereas there was no significant difference in the plasma level of these proteins between the groups on Day 0 (Dataset S9).

**Figure 6.**
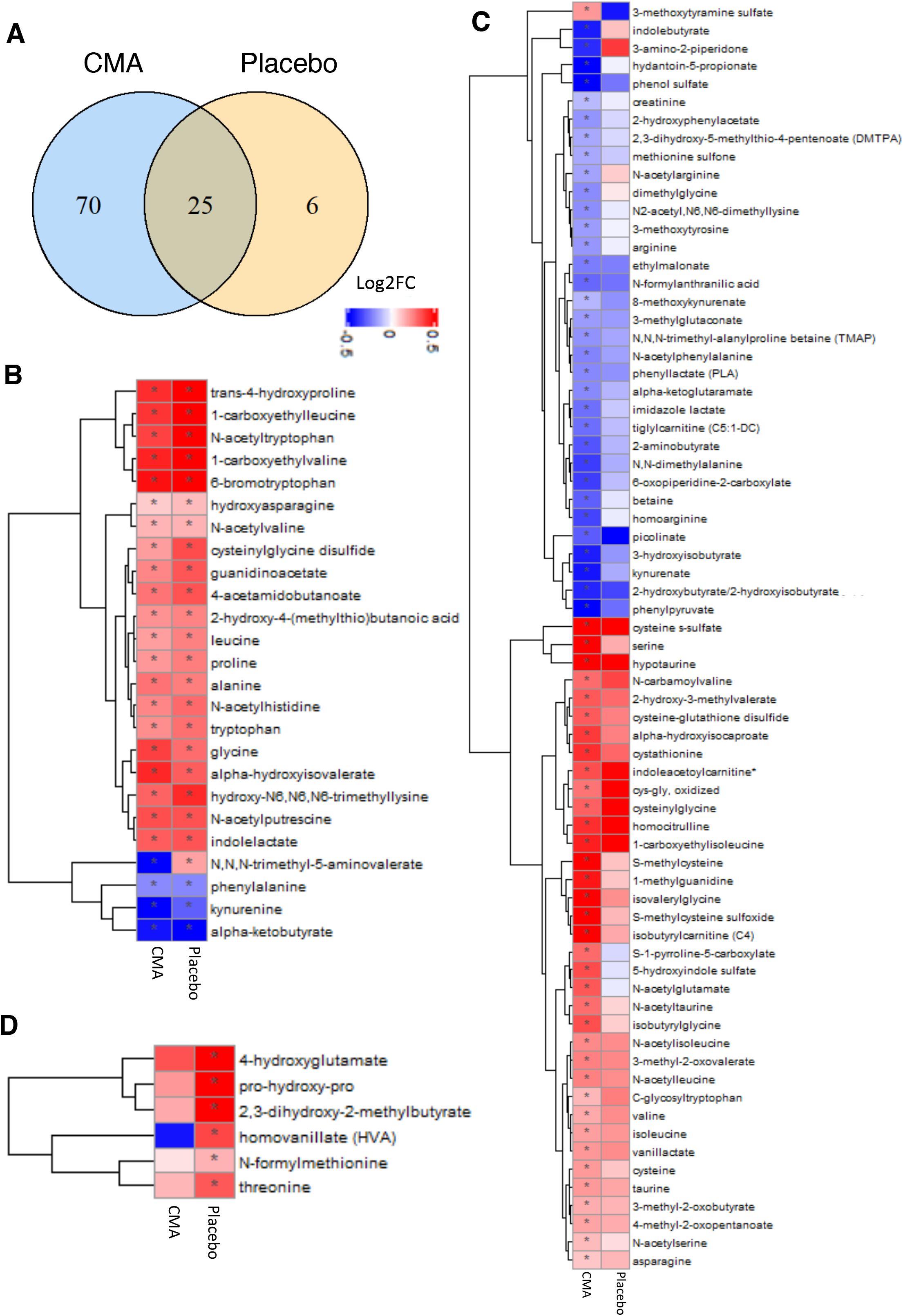
CMA effect the plasma level of amino acids and improve metabolic health. Plasma level of amino acids that are significantly different on Days 14 vs Day 0 in the CMA and placebo groups. **A)** Venn-diagram representing the number of identified amino acids that are significantly different on Days 14 vs Day 0 in the CMA and placebo groups. The intersection represents amino acids that were altered in both groups. Statistical significance based on paired Student’s t test. FDR value: < 0.001. Association between the plasma level of significantly different amino acids **B)** in both groups (n=25); **C)** only in CMA group (n=70) and **D)** only in placebo group (n=6) on Day 14 vs Day 0. Heatmap shows log2FC based alterations in the amino acids and asterisks indicate statistical significance based on paired Student’s t test. FDR value: < 0.001. Log2FC, log2(fold change).

We investigated the association between the plasma cytokine levels and the individual metabolic activators and found that the plasma level of most cytokine levels were significantly inversely correlated with the plasma level of serine (Figure 5B, Dataset S10). Hence, based on our analysis, we suggest that serine is a key metabolic activator leading to the decrease in the cytokine levels and improvement in metabolic conditions in COVID-19 patients.

### 2.8 Effect of CMA on Global Metabolism

We identified the significantly (FDR<0.001) different plasma metabolites on Day 14 vs Day 0 in both CMA and placebo groups. We found that the plasma levels of 122 metabolites were significantly different in both CMA and placebo groups, whereas plasma levels of 231 and 20 metabolites were specifically significantly different only in the CMA and placebo groups, respectively (FDR<0.001, Dataset S4). Evaluation of plasma metabolites that differed significantly on Day 14 vs Day 0 in each group showed that larger number of metabolites related to amino acid (Figure 6A), lipid metabolism (Figure S5) and other metabolic pathways (Figure S6) were altered in CMA group compared to the placebo group (Dataset S4). By investigating the metabolites involved in amino acid metabolism, we found that plasma levels of 25 metabolites were significantly different both in the CMA and placebo group, whereas 70 and 6 were specifically significantly altered only in the CMA and placebo group, respectively (FDR<0.001, Figure 6A, Dataset S4).

Increased plasma levels of metabolites in the kynurenine pathway were associated with COVID-19 severity (*31*). In our study, we found that plasma levels of kynurenate and 8-methoxykynurenate were significantly lower on Day 14 vs Day 0 in the CMA group (Figure 6B, Dataset S4). The plasma level of kynurenine was significantly decreased on Day 14 vs Day 0 both in the CMA and placebo groups. However, the magnitude of the reduction of kynurenine was significant in the CMA vs placebo on Day 14 (Figure 6C, Dataset S4 & S5). Reduction in the plasma level of kynurenine and kynurenate was positively correlated with plasma serine (Dataset S6).

Plasma levels of metabolites related to the urea cycle (3-amino-2-piperidone, N,N,N-trimethyl-alanyl proline betaine, homoarginine, and arginine) were significantly decreased in the CMA vs placebo group on Day 14 (Figure 6B, Dataset S4). Specifically, the plasma level of 3-amino-2-piperidone was significantly decreased in CMA group on Day 14 vs Day 0 (Figure 6B, Dataset S4) and inversely correlated with the plasma level of serine (Dataset S6).

Emerging evidence indicates that kidney complications in COVID-19 patients are frequent, but the potential impact of SARS-Cov2 on the kidney is still undetermined (*32*). Recent studies showed that plasma level of N,N,N-trimethyl-5-aminovalerate involved in lysine metabolism is an indicator of elevated urinary albumin excretion (*33*). In our study, we found that the plasma level of N,N,N-trimethyl-5-aminovalerate was significantly increased on Day 14 vs Day 0 in the placebo group while its plasma level is significantly decreased on Day 14 vs Day 0 in the CMA group (Figure 6B, Dataset S4). Moreover, the plasma level of creatinine was only significantly decreased on Day 14 vs Day 0 in the CMA group (Figure 6B, Dataset S4). Plasma level of both creatinine and N,N,N-trimethyl-5-aminovalerate were significantly decreased in the CMA vs placebo group on Day 14 (Dataset S5). The plasma reduction on N,N,N-trimethyl-5-aminovalerate is significantly inversely correlated with the plasma level of serine (Dataset S6).

Lipids play a fundamental role in multiple stages of the virus cycle. Specific lipid species of cellular membranes (e.g. cholesterol and sphingolipids) are not only barriers but also receptors to infection initiation (*34*). In our study, plasma level of 71 metabolites involved in lipid metabolism were significantly (FDR<0.001) different in both groups, whereas 102 and 5 were specifically significantly changed only in the CMA and placebo groups, respectively (Figure S5A, Dataset S4). Plasma level of a significant number of metabolites associated with ceramides and sphingomyelins were significantly decreased on Day 14 vs Day 0 in both CMA and placebo groups (Figure S5B, Dataset S4). However, the reduction in the plasma level of metabolites involved in ceramides and sphingomyelins was more dramatic in the CMA group (Figure S5B, Dataset S4). Plasma level of metabolites associated with fatty acid metabolism (acyl carnitines) were significantly increased in the CMA group on Day 14 vs Day 0. In both groups, plasma levels of lysophospholipid, phosphatidylcholine, and androgenic steroids were significantly increased on Day 14 vs Day 0 (Figure S5B and C, Dataset S4). These alterations were significantly positively correlated with carnitine and serine levels (Dataset S6).

Ketone bodies have important signaling roles in restoring altered energy metabolism and redox state. In this study, we found that the plasma level of 2R,3R-dihydroxybutyrate was significantly decreased in the CMA group on Day 14 vs Day 0 (Figure S5B, Dataset S4). We also observed that plasma level of carnitine and serine also showed a significant inverse correlation with the plasma level of 2R,3R-dihydroxybutyrate (Dataset S6). We also found that vitamins (Tocopherol, vitamin A, and riboflavin) and benzoate metabolism were significantly increased in both groups, whereas the plasma level of metabolites associated with the TCA cycle intermediate products were upregulated on Day 14 vs Day 0 in the CMA (Figure S6, Dataset S4). Overall, these results indicate that the administration of CMA effected the global metabolism of the patients.

### 2.9 Integrative Analysis of Clinical Variables with Proteomics and Metabolomics data

We have studied the associations between the significantly improved (decreased) clinical parameters including AST, ALT, LDH and TGs with the plasma level of all metabolites (Dataset S11) and all inflammation proteins (Dataset S12) and presented the 10 most significantly correlated metabolites (Figure 7A) and inflammation proteins (Figure 7B). The reduction in all four clinical parameters was significantly correlated with the plasma level of serine and glycine. We also found that plasma level of cysteine-glutathione disulfide was significantly inversely correlated with the level of AST ALT and TGs. Hence, we propose that serine is the key metabolic activator leading to the improvement in the clinical parameters in COVID-19 patients.

**Figure 7.**
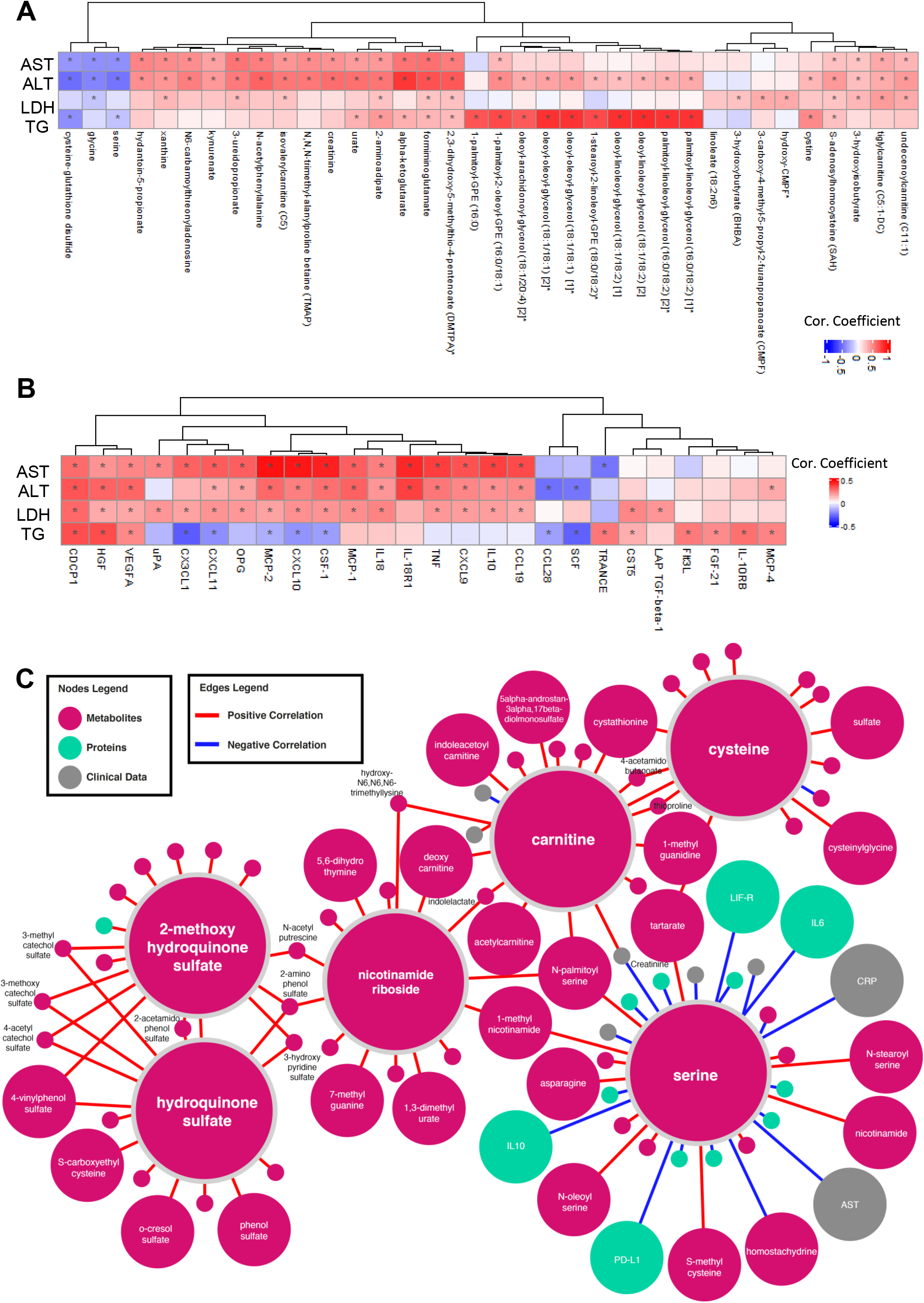
Integrated network analysis for revealing the effect of CMA on patients. Association between the plasma level of clinical variables including ALT, AST, LDH, TGs and hemoglobin with plasma level of all inflammation related proteins. Heatmap shows log2FC based alterations and clustering between the plasma level of clinical variables including ALT, AST, LDH, TG and hemoglobin with plasma level of **A)** all metabolites and **B)** all inflammation related proteins. Asterisks indicate statistical significance based on paired Student’s t test. p value: < 0.05. Log2FC, log2(fold change); AST, Aspartate aminotransferase; ALT, Alanine transaminase; LDH, Lactate dehydrogenase; TGs, Triglycerides. **C)** Neighbors of the metabolic activators including serine, carnitine, nicotinamide riboside, and cysteine as well as degradation products of hydroxychloroquine including hydroquinone sulfate and 2-methoxyhydroquinone sulfate (1) based on the multi-omics network analysis. Only analytes that are significantly altered in CMA Day 14 vs Day 0 are highlighted. Full networks can be found in iNetModels (http://inetmodels.com).

We also evaluated the association of plasma proteins with the level of AST, ALT, LDH and TG and found that CDCP1 was positively correlated with all parameters (Figure 7B, Dataset S12). Plasma level of a set of inflammatory proteins (MCP-2, CXCL10, CSF-1, IL-18R1, TNF, MCP-1, CDCP1 and VEGFA) were significantly positively correlated with the level of ALT and AST (Figure 7B). Interestingly, we found that plasma levels of IL10, CCL19, CXCL9, CXCL1, OPG, CX3CL1, PD-L1, LIF-R, IFN-gamma and TRAIL were positively correlated and TRANCE, IL18, IL-12B, TWEAK, uPA and IL6 were inversely correlated with the level of AST, however these proteins were not significantly correlated with the level of ALT. On the other hand, the level of ALT was also significantly positively correlated with HGF and negatively correlated with CCL28 and SCF, which were not significantly correlated with the level of AST. Additionally, level of TG was positively correlated with the plasma level of TRANCE, IL-10RB, CDCP1 and HGF, however its inverse correlation was observed with the plasma level of CX3CL1 and SCF. We also found that hemoglobin levels were significantly correlated with the inflammatory proteins including EN-RAGE, DNER, IL-10RB, VEGFA, HGF, CST5 and MCP-1 (Dataset S12).

Finally, we constructed a multi-omics correlation network based on clinical variables, proteomics, and metabolomics data (Dataset S13, unfiltered network). The full network can be explored via iNetModels (http://inetmodels.com), an open-access interactive web platform for multi-omics data visualization and database. This network showed direct and indirect intra- and inter-omics functional relationships of different analytes. We subsequently overlaid the statistically altered analytes (clinical variables, metabolites and proteins) to the sub-network of the individual metabolic activators and degradation products of HQ (Figure 7C). The subnetwork was built from the union of <u>top 15 metabolite neighbors</u>, and <u>all protein and clinical neighbors</u>. We identified the serine as the most connected metabolic activator to clinical variables, metabolites, and proteins. Moreover, we performed centrality analysis of the network to identify the most central inflammation protein markers, and found that MCP-4, IL-18R1, HGF, CXCL10 and CSF-1 were among top 10 most-central (degree) protein nodes which all showed significant downregulation on Day 14 vs Day 0 in the CMA group. We found that all of them, except MCP-4, were significantly inversely correlation with serine (Dataset S14). Hence, based on multi-omics network analysis, we confirmed our findings based on single-omics analysis and propose that serine acts as the key metabolic activator in the CMA and the effect seen in the clinical study is mostly due to the serine in the CMA.

## 3. CONCLUSION AND DISCUSSION

Our study shows that the combination of CMA and standard therapy significantly reduces the mean recovery time of ambulatory patients with COVID-19 as compared with placebo and standard therapy (6.6 vs 9.3 days in phase-2) and (5.7 vs 9.2 days in phase-3). Our multi-omics analysis on patients recruited in phase-2 trial is consistent with the notion that CMA improves clinical outcomes in COVID-19 by improving immune response, and regulating antioxidant, amino acid and lipid metabolism. After having a positive result on the phase 2 study with 93 patients, we designed a double-blinded placebo-controlled Phase 3 study with 304 patients. We have shown that administration of the CMA accelerates the recovery of the patients independent of standard therapy. We have also shown that the clinical parameters including ALT, LDH, creatinine and glucose were significantly improved after the administration of the CMA in a total of 397 patients recruited in phase 2 and phase 3 trial. In the phase 3 trial, we also included the patients treated with FP, and again showed that CMA accelerates the recovery of the patients independent of the standard therapy. We also compared the effect of the HQ and FP in the recovery of the patients and observed no significant differences between the drugs used in the standard therapy.

Although COVID-19 primarily causes symptoms of respiratory infection, it often causes gastrointestinal and hepatic symptoms, including nausea/vomiting, diarrhea, and elevated levels of liver enzymes (*1*). Some studies have reported that liver deficiencies correlate with worse outcomes, including longer hospitalization, progression to severe COVID-19, admission to the intensive care unit, and death (*35-38*). Mounting evidence shows that the GSH level is insufficient to maintain and regulate the thiol redox status of the liver in subjects with liver dysfunction (*26*). Depleted liver GSH can be restored by administration of NAC, serine and glycine which can be synthesized through the interconversion of serine. Serine synthesis is downregulated in NAFLD patients, and administration of serine enhances homocysteine metabolism in mice and rats (*39*). Two other components of CMA including carnitine and NR, which stimulate the transfer of fatty acids from the cytosol to mitochondria, are also depleted in liver diseases (*40-43*). Hence, we have here analyzed the effect of a mixture of CMA consisting of serine, a cysteine analogue, salt form of L-carnitine and an NAD+ precursor, administrated to COVID-19 patients to increase the GSH level in liver and other tissues.

NAD^+^ is an essential metabolite in cellular energy generation and redox biology. Recently, it has been shown that the anti-coronaviral activities of four noncanonical PARP isozyme activities are limited by cellular NAD^+^ status (*44*). Additionally, a significant activation of kynurenine pathway in severe COVID-19 population has been reported (*31*). Tryptophan is an essential amino acid that is fundamental for the biosynthesis of fundamental neuromodulators, namely serotonin and melatonin. Tryptophan catabolism through kynurenine pathway is the sole route for de novo NAD+ synthesis. In this context, CMA significantly decreased the plasma levels of metabolites involved in kynurenine pathway, and this reduction might play a crucial role in restoring the metabolic balance in COVID-19 patients.

As with all metabolomics studies, not every metabolite can be captured in a single assay. In this study, we were unable to measure plasma NAD+ in the study participants. NAD+ levels in the plasma are dynamic and are typically much lower than levels found in the cells and tissues. Unfortunately, plasma NR is not a surrogate marker for NAD+ status and has an estimated half-life of 2.7 hours and a C_max_ of 3 hours(*45*). In a clinical kinetics study following 9 days of NR supplementation, NR levels still appeared to peak and trough at 3 hours, however NAD+ levels were consistent across time points, having reached steady state (*45*). Any correlations related to plasma NR and the up- or down-regulation of metabolites or inflammatory markers is not likely reflective of actual NAD+ status in the study participants. Future research that evaluates NAD+ levels in COVID-19 patients before and after CMA treatment, as well as treatment with the individual co-factors separately would provide further evidence of the role of NAD+ status on potential correlation (and causation) to changes in the metabolome and inflammatory markers.

In this study, patients also received HQ or FP, recommended as standard therapy in some countries. Even though, we assessed the associations between the plasma level of HQ products and other metabolites, we could not rule out possible drug interactions based on existing data. Based on metabolomics data, we observed that N-acetyltryptophan is significantly positively correlated with HQ products and the plasma level of N-acetyltryptophan is increased more in placebo group (log2FC=0.60) than CMA group (log2FC=0.36). Unfortunately, due to access limitations to the patients during the study, we were unable to sample the patients directly following the end of HQ or FP treatment. An additional mid-point analysis at 7 days may have provided additional data related to the HQ metabolites in response to the inclusion of CMA.

Many COVID-19 patients are at risk for detrimental outcomes due to systemic inflammatory responses referred to as a cytokine storm, a life-threatening condition is dependent on downstream processes that lead to oxidative stress, dysregulation of iron homeostasis, hypercoagulability, and thrombocytopenia (*46, 47*). Several studies have proposed that CMA components may effectively inhibit the production of proinflammatory molecules (e.g., IL-6, CCL-5, CXCL-8, and CXCL-10) and improve impaired mitochondrial functions by reducing oxidative damage, lipid peroxidation, and disturbed glucose tolerance (*44, 48*). Based on integrative analysis, we observed that CMA may interrupt the overactive immune response and early treatment with CMA may reduce the risk of progression to severe respiratory distress and lung damage.

In conclusion, we evaluated the efficacy and safety of CMA in combination with HQ or FP therapy in patients with mild-to-moderate COVID-19 and observed that combination therapy is safe and beneficial in patients with mild-to-moderate COVID-19. Our analysis suggests that CMA is am effective treatment for COVID-19.

### 4.4 EXPERIMENTAL SECTION

#### Trial Design and Oversight

Patients for this randomized, open-label, placebo-controlled, phase-2 study and randomized, open-label, double-blinded, phase-3 study were recruited at the Umraniye Training and Research Hospital, University of Health Sciences, Istanbul, Turkey. Written informed consent was obtained from all participants before the initiation of any trial-related procedures. The safety of the participants and the risk–benefit analysis was overseen by an independent external data-monitoring committee. The trial was conducted in accordance with Good Clinical Practice guidelines and the principles of the Declaration of Helsinki. The study was approved by the ethics committee of Istanbul Medipol University, Istanbul, Turkey, and retrospectively registered at https://clinicaltrials.gov/with (Clinical Trial ID: NCT04573153).

#### Participants

Patients were enrolled in the trial if they were over 18 years of age, had a PCR-confirmed COVID-19 test within the previous 24 h, and were in stable condition not requiring hospitalization. Chest tomography was done to rule out pneumonia. Patients who had a partial oxygen saturation below 93% and required hospitalization after diagnosis were excluded. The main characteristics of the patients are summarized in Table S2. The inclusion, exclusion, and randomization criteria are described in detail in the Supplementary Appendix.

#### Randomization, Interventions, and Follow-up

Patients were randomly assigned to receive CMA or placebo (3:1) and also received standard therapy with hydroxychloroquine or FP. Patient information (patient number, date of birth, initials) was entered into the web-based randomization system, and the randomization codes were entered into the electronic case report form. Except the investigator, who had password access to the system, all other clinical staff were blinded to treatment, as were the participants.

Treatment started on the day of diagnosis. Both placebo and CMA were provided in powdered form in identical plastic bottle containing a single dose to be dissolved in water and taken orally one dose in the morning after breakfast and one dose in the evening after dinner. Each dose of CMA contained 3.73 g L-carnitine tartrate, 2.55 g N-acetylcysteine, 1 g nicotinamide riboside chloride, and 12.35 g serine. All participants also received oral HQ or FP for 5 days. The patients were contacted by telephone daily to assess symptoms and adverse events. All patients came for a follow-up visit on Day 14. Further information is provided in the Supplementary Appendix.

#### Outcomes

The primary end point in the original protocol was to assess the clinical efficacy of the combination of CMA in COVID-19 patients. For the primary purpose, the proportion of patients who fully recovered from COVID-19, as demonstrated by being symptom free within the 14 days of the initial diagnosis of COVID-19 was determined This was amended to include self-reporting of daily symptoms and clinical status using a binomial scale (present/absent) via daily telephone visits by clinical staff. The secondary aim in this study was to evaluate the safety and tolerability of CMA and HQ or FP combination. All protocol amendments were authorized and approved by the sponsor, the institutional review board or independent ethics committee, and the pertinent regulatory authorities.

Number and characteristics of adverse events, serious adverse events, and treatment discontinuation due to CMA were reported from the beginning of the study to the end of the follow-up period as key safety endpoints. The changes in vital signs (systolic and diastolic blood pressures, pulse, respiratory rate, body temperature, pulse oximetry values), baseline values, and the status of treatment were recorded at day 0 and 14. A complete list of the end points is provided in the Supplementary Appendix.

#### Inflammatory Proteomics

Plasma levels of inflammatory proteins were determined with the Olink Inflammation panel (Olink Bioscience, Uppsala, Sweden). Briefly each sample was incubated with 92 DNA-labeled antibody pairs (proximity probes). When an antibody pair binds to its corresponding antigens, the corresponding DNA tails form an amplicon by proximity extension, which can be quantified by high-throughput, real-time PCR. Probe solution (3 μl) was mixed with 1 μl of sample and incubated overnight at 4°C. Then 96 μl of extension solution containing extension enzyme and PCR reagents for the pre-amplification step was added, and the extension products were mixed with detection reagents and primers and loaded on the chip for qPCR analysis with the BioMark HD System (Fluidigm Corporation, South San Francisco, CA).To minimize inter- and intrarun variation, the data were normalized to both an internal control and an interplate control. Normalized data were expressed in arbitrary units (Normalized Protein eXpression, NPX) on a log2 scale and linearized with the formula 2^NPX^. A high NPX indicates a high protein concentration. The limit of detection, determined for each of the 92 assays, was defined as three standard deviations above the negative control (background).

#### Untargeted Metabolomics Analysis

Plasma samples were collected on Days 0 and 14 for nontargeted metabolite profiling by Metabolon (Durham, NC). The samples were prepared with an automated system (MicroLab STAR, Hamilton Company, Reno, NV). For quality control purposes, a recovery standard was added before the first step of the extraction. To remove protein and dissociated small molecules bound to protein or trapped in the precipitated protein matrix, and to recover chemically diverse metabolites, proteins were precipitated with methanol under vigorous shaking for 2 min (Glen Mills GenoGrinder 2000) and centrifuged. The resulting extract was divided into four fractions: one each for analysis by ultraperformance liquid chromatography–tandem mass spectroscopy (UPLC-MS/MS) with positive ion-mode electrospray ionization, UPLC-MS/MS with negative ion-mode electrospray ionization, and gas chromatography–mass spectrometry; one fraction was reserved as a backup.

#### Statistical Analysis

The primary outcome was the time to recovery, defined as the time from diagnosis to the first self-report of no symptom. Recovery curves were generated with the Kaplan-Meier method. Hazard ratios and 95% confidence intervals were calculated with the Univariate and Multivariate Cox proportional-hazards regression model based on R package survival. We adjusted the age, gender, other treatment history, cigarette, alcohol and HQ/FP treatment in Multivariate Cox regression analysis. In phase-2 study, we excluded the HQ/FP factor since all the patients received HQ treatment and none of them received FP treatment. F

Differences in clinical measurements, proteomics, and metabolomics data were analyzed by t-test (paired t-test within group comparison). The correlations for multi-omics networks were calculated using Spearman correlation and filtered with FDR < 0.05. The analyses were performed with SciPy package in Python 3.7. Centrality analysis on network was performed using Cytoscape 3.8.2. The script used to analyze the data can be found at https://github.com/sysmedicine/AltayEtAll_2020_Covid_CMA

## Data Availability

The data that support the findings of this study are available from the corresponding author, upon reasonable request.

## ACKNOWLEDGMENTS

This work was financially supported by ScandiBio Therapeutics and Knut and Alice Wallenberg Foundation. The authors would like to thank to the Plasma Profiling Facility team at SciLifeLab in Stockholm for generating the Olink data, Metabolon Inc. (Durham, USA) for generation of metabolomics data, and ChromaDex Inc. (Irvine, CA, USA) for providing NR. AM and HY acknowledge support from the PoLiMeR Innovative Training Network (Marie Sk∤odowska-Curie Grant Agreement No. 812616) which has received funding from the European Union’s Horizon 2020 research and innovation programme.

## SUPPORTING INFORMATION

Supporting Information includes Supplemental Appendix, 6 Supplementary Figures and 14 Supplementary Datasets.

## REFERENCES

1. A. Gupta, M. V. Madhavan, K. Sehgal, N. Nair, S. Mahajan, T. S. Sehrawat, B. Bikdeli, N. Ahluwalia, J. C. Ausiello, E. Y. Wan, D. E. Freedberg, A. J. Kirtane, S. A. Parikh, M. S. Maurer, A. S. Nordvig, D. Accili, J. M. Bathon, S. Mohan, K. A. Bauer, M. B. Leon, H. M. Krumholz, N. Uriel, M. R. Mehra, M. S. V. Elkind, G. W. Stone, A. Schwartz, D. D. Ho, J. P. Bilezikian, D. W. Landry, Extrapulmonary manifestations of COVID-19. Nature Medicine 26, 1017–1032 (2020).

2. Johns Hopkins University of Medicine. (2021, January 11).

3. L. Zhu, Z.-G. She, X. Cheng, J.-J. Qin, X.-J. Zhang, J. Cai, F. Lei, H. Wang, J. Xie, W. Wang, H. Li, P. Zhang, X. Song, X. Chen, M. Xiang, C. Zhang, L. Bai, D. Xiang, M.-M. Chen, Y. Liu, Y. Yan, M. Liu, W. Mao, J. Zou, L. Liu, G. Chen, P. Luo, B. Xiao, C. Zhang, Z. Zhang, Z. Lu, J. Wang, H. Lu, X. Xia, D. Wang, X. Liao, G. Peng, P. Ye, J. Yang, Y. Yuan, X. Huang, J. Guo, B.-H. Zhang, H. Li, Association of Blood Glucose Control and Outcomes in Patients with COVID-19 and Pre-existing Type 2 Diabetes. Cell Metabolism 31, 1068-1077.e1063 (2020).

4. J.-W. Song, S. M. Lam, X. Fan, W.-J. Cao, S.-Y. Wang, H. Tian, G. H. Chua, C. Zhang, F.-P. Meng, Z. Xu, J.-L. Fu, L. Huang, P. Xia, T. Yang, S. Zhang, B. Li, T.-J. Jiang, R. Wang, Z. Wang, M. Shi, J.-Y. Zhang, F.-S. Wang, G. Shui, Omics-Driven Systems Interrogation of Metabolic Dysregulation in COVID-19 Pathogenesis. Cell Metabolism 32, 188-202.e185 (2020).

5. T. Thomas, D. Stefanoni, J. A. Reisz, T. Nemkov, L. Bertolone, R. O. Francis, K. E. Hudson, J. C. Zimring, K. C. Hansen, E. A. Hod, S. L. Spitalnik, A. D’Alessandro, COVID-19 infection alters kynurenine and fatty acid metabolism, correlating with IL-6 levels and renal status. JCI Insight 5, (2020).

6. S. R. Bornstein, R. Dalan, D. Hopkins, G. Mingrone, B. O. Boehm, Endocrine and metabolic link to coronavirus infection. Nature Reviews Endocrinology 16, 297–298 (2020).

7. F. Zhou, T. Yu, R. Du, G. Fan, Y. Liu, Z. Liu, J. Xiang, Y. Wang, B. Song, X. Gu, L. Guan, Y. Wei, H. Li, X. Wu, J. Xu, S. Tu, Y. Zhang, H. Chen, B. Cao, Clinical course and risk factors for mortality of adult inpatients with COVID-19 in Wuhan, China: a retrospective cohort study. The Lancet 395, 1054–1062 (2020).

8. I. F.-N. Hung, K.-C. Lung, E. Y.-K. Tso, R. Liu, T. W.-H. Chung, M.-Y. Chu, Y.-Y. Ng, J. Lo, J. Chan, A. R. Tam, H.-P. Shum, V. Chan, A. K.-L. Wu, K.-M. Sin, W.-S. Leung, W.-L. Law, D. C. Lung, S. Sin, P. Yeung, C. C.-Y. Yip, R. R. Zhang, A. Y.-F. Fung, E. Y.-W. Yan, K.-H. Leung, J. D. Ip, A. W.-H. Chu, W.-M. Chan, A. C.-K. Ng, R. Lee, K. Fung, A. Yeung, T.-C. Wu, J. W.-M. Chan, W.-W. Yan, W.-M. Chan, J. F.-W. Chan, A. K.-W. Lie, O. T.-Y. Tsang, V. C.-C. Cheng, T.-L. Que, C.-S. Lau, K.-H. Chan, K. K.-W. To, K.-Y. Yuen, Triple combination of interferon beta-1b, lopinavir–ritonavir, and ribavirin in the treatment of patients admitted to hospital with COVID-19: an open-label, randomised, phase 2 trial. The Lancet 395, 1695–1704 (2020).

9. O. Altay, E. Mohammadi, S. Lam, H. Turkez, J. Boren, J. Nielsen, M. Uhlen, A. Mardinoglu, Current Status of COVID-19 Therapies and Drug Repositioning Applications. iScience 23, 101303 (2020).

10. The U.S. Food and Drug Administration (FDA) (2020, August 28).

11. University of South Carolina, Regulatory updates on China, February 2020. 2020, September 18).

12. R. Miller, A. R. Wentzel, G. A. Richards, COVID-19: NAD(+) deficiency may predispose the aged, obese and type2 diabetics to mortality through its effect on SIRT1 activity. Med Hypotheses 144, 110044–110044 (2020).

13. F. Silvagno, A. Vernone, G. P. Pescarmona, The Role of Glutathione in Protecting against the Severe Inflammatory Response Triggered by COVID-19. Antioxidants (Basel) 9, 624 (2020).

14. E. H. Ma, G. Bantug, T. Griss, S. Condotta, R. M. Johnson, B. Samborska, N. Mainolfi, V. Suri, H. Guak, M. L. Balmer, M. J. Verway, T. C. Raissi, H. Tsui, G. Boukhaled, S. Henriques da Costa, C. Frezza, C. M. Krawczyk, A. Friedman, M. Manfredi, M. J. Richer, C. Hess, R. G. Jones, Serine Is an Essential Metabolite for Effector T Cell Expansion. Cell Metabolism 25, 345–357 (2017).

15. M. Mata, I. Sarrion, M. Armengot, C. Carda, I. Martinez, J. A. Melero, J. Cortijo, Respiratory syncytial virus inhibits ciliagenesis in differentiated normal human bronchial epithelial cells: effectiveness of N-acetylcysteine. PLoS One 7, e48037–e48037 (2012).

16. Q. Zhang, Y. Ju, Y. Ma, T. Wang, N-acetylcysteine improves oxidative stress and inflammatory response in patients with community acquired pneumonia: A randomized controlled trial. Medicine (Baltimore) 97, e13087–e13087 (2018).

17. Y. Zhang, S. Ding, C. Li, Y. Wang, Z. Chen, Z. Wang, Effects of N-acetylcysteine treatment in acute respiratory distress syndrome: A meta-analysis. Exp Ther Med 14, 2863–2868 (2017).

18. S. Sharma, A. Aramburo, R. Rafikov, X. Sun, S. Kumar, P. E. Oishi, S. A. Datar, G. Raff, K. Xoinis, G. Kalkan, S. Fratz, J. R. Fineman, S. M. Black, L-Carnitine preserves endothelial function in a lamb model of increased pulmonary blood flow. Pediatric Research 74, 39–47 (2013).

19. G. Malaguarnera, M. Pennisi, C. Gagliano, M. Vacante, M. Malaguarnera, S. Salomone, F. Drago, G. Bertino, F. Caraci, G. Nunnari, M. Malaguarnera, Acetyl-L-Carnitine Supplementation During HCV Therapy With Pegylated Interferon-α 2b Plus Ribavirin: Effect on Work Performance; A Randomized Clinical Trial. Hepat Mon 14, e11608–e11608 (2014).

20. A. Balasubramanyam, I. Coraza, E. O. Smith, L. W. Scott, P. Patel, D. Iyer, A. A. Taylor, T. P. Giordano, R. V. Sekhar, P. Clark, E. Cuevas-Sanchez, S. Kamble, C. M. Ballantyne, H. J. Pownall, Combination of niacin and fenofibrate with lifestyle changes improves dyslipidemia and hypoadiponectinemia in HIV patients on antiretroviral therapy: results of “heart positive,” a randomized, controlled trial. J Clin Endocrinol Metab 96, 2236–2247 (2011).

21. A. E. Rodriguez, G. S. Ducker, L. K. Billingham, C. A. Martinez, N. Mainolfi, V. Suri, A. Friedman, M. G. Manfredi, S. E. Weinberg, J. D. Rabinowitz, N. S. Chandel, Serine Metabolism Supports Macrophage IL-1β Production. Cell Metabolism 29, 1003-1011.e1004 (2019).

22. X. Zhou, H. Zhang, L. He, X. Wu, Y. Yin, Long-Term l-Serine Administration Reduces Food Intake and Improves Oxidative Stress and Sirt1/NFκB Signaling in the Hypothalamus of Aging Mice. Front Endocrinol (Lausanne) 9, 476 (2018).

23. D. Conze, C. Brenner, C. L. Kruger, Safety and Metabolism of Long-term Administration of NIAGEN (Nicotinamide Riboside Chloride) in a Randomized, Double-Blind, Placebo-controlled Clinical Trial of Healthy Overweight Adults. Sci Rep 9, 9772 (2019).

24. A. Mardinoglu, D. Ural, M. Zeybel, H. H. Yuksel, M. Uhlén, J. Borén, The Potential Use of Metabolic Cofactors in Treatment of NAFLD. Nutrients 11, 1578 (2019).

25. S. A. J. Trammell, M. S. Schmidt, B. J. Weidemann, P. Redpath, F. Jaksch, R. W. Dellinger, Z. Li, E. D. Abel, M. E. Migaud, C. Brenner, Nicotinamide riboside is uniquely and orally bioavailable in mice and humans. Nature Communications 7, 12948 (2016).

26. A. Mardinoglu, E. Bjornson, C. Zhang, M. Klevstig, S. Soderlund, M. Stahlman, M. Adiels, A. Hakkarainen, N. Lundbom, M. Kilicarslan, B. M. Hallstrom, J. Lundbom, B. Verges, P. H. Barrett, G. F. Watts, M. J. Serlie, J. Nielsen, M. Uhlen, U. Smith, H. U. Marschall, M. R. Taskinen, J. Boren, Personal model-assisted identification of NAD(+) and glutathione metabolism as intervention target in NAFLD. Mol Syst Biol 13, 916 (2017).

27. Y. S. Elhassan, K. Kluckova, R. S. Fletcher, M. S. Schmidt, A. Garten, C. L. Doig, D. M. Cartwright, L. Oakey, C. V. Burley, N. Jenkinson, M. Wilson, S. J. E. Lucas, I. Akerman, A. Seabright, Y. C. Lai, D. A. Tennant, P. Nightingale, G. A. Wallis, K. N. Manolopoulos, C. Brenner, A. Philp, G. G. Lavery, Nicotinamide Riboside Augments the Aged Human Skeletal Muscle NAD(+) Metabolome and Induces Transcriptomic and Anti-inflammatory Signatures. Cell Rep 28, 1717-1728.e1716 (2019).

28. A. Mardinoglu, J. Boren, U. Smith, M. Uhlen, J. Nielsen, Systems biology in hepatology: approaches and applications. Nature Reviews Gastroenterology & Hepatology 15, 365–377 (2018).

29. C. Zhang, E. Bjornson, M. Arif, A. Tebani, A. Lovric, R. Benfeitas, M. Ozcan, K. Juszczak, W. Kim, J. T. Kim, G. Bidkhori, M. Ståhlman, P.-O. Bergh, M. Adiels, H. Turkez, M.-R. Taskinen, J. Bosley, H.-U. Marschall, J. Nielsen, M. Uhlén, J. Borén, A. Mardinoglu, The acute effect of metabolic cofactor supplementation: a potential therapeutic strategy against non-alcoholic fatty liver disease. Mol Syst Biol 16, e9495–e9495 (2020).

30. F. Coperchini, L. Chiovato, L. Croce, F. Magri, M. Rotondi, The cytokine storm in COVID-19: An overview of the involvement of the chemokine/chemokine-receptor system. Cytokine Growth Factor Rev 53, 25–32 (2020).

31. B. Shen, X. Yi, Y. Sun, X. Bi, J. Du, C. Zhang, S. Quan, F. Zhang, R. Sun, L. Qian, W. Ge, W. Liu, S. Liang, H. Chen, Y. Zhang, J. Li, J. Xu, Z. He, B. Chen, J. Wang, H. Yan, Y. Zheng, D. Wang, J. Zhu, Z. Kong, Z. Kang, X. Liang, X. Ding, G. Ruan, N. Xiang, X. Cai, H. Gao, L. Li, S. Li, Q. Xiao, T. Lu, Y. Zhu, H. Liu, H. Chen, T. Guo, Proteomic and Metabolomic Characterization of COVID-19 Patient Sera. Cell 182, 59-72.e15 (2020).

32. C. Benedetti, M. Waldman, G. Zaza, L. V. Riella, P. Cravedi, COVID-19 and the Kidneys: An Update. Front Med (Lausanne) 7, 423 (2020).

33. J. K. Haukka, N. Sandholm, C. Forsblom, J. E. Cobb, P.-H. Groop, E. Ferrannini, Metabolomic Profile Predicts Development of Microalbuminuria in Individuals with Type 1 Diabetes. Scientific Reports 8, 13853 (2018).

34. N. S. Heaton, G. Randall, Multifaceted roles for lipids in viral infection. Trends Microbiol 19, 368–375 (2011).

35. M. A. Hundt, Y. Deng, M. M. Ciarleglio, M. H. Nathanson, J. K. Lim, Abnormal Liver Tests in COVID-19: A Retrospective Observational Cohort Study of 1827 Patients in a Major U.S. Hospital Network. Hepatology, (2020).

36. K. P. Patel, P. A. Patel, R. R. Vunnam, A. T. Hewlett, R. Jain, R. Jing, S. R. Vunnam, Gastrointestinal, hepatobiliary, and pancreatic manifestations of COVID-19. J Clin Virol 128, 104386 (2020).

37. W. D. Redd, J. C. Zhou, K. E. Hathorn, T. R. McCarty, A. N. Bazarbashi, C. C. Thompson, L. Shen, W. W. Chan, Prevalence and Characteristics of Gastrointestinal Symptoms in Patients With Severe Acute Respiratory Syndrome Coronavirus 2 Infection in the United States: A Multicenter Cohort Study. Gastroenterology 159, 765-767.e762 (2020).

38. A. Bertolini, I. P. van de Peppel, F. A. J. A. Bodewes, H. Moshage, A. Fantin, F. Farinati, R. Fiorotto, J. W. Jonker, M. Strazzabosco, H. J. Verkade, G. Peserico, Abnormal liver function tests in COVID-19 patients: relevance and potential pathogenesis. Hepatology n/a.

39. W.-C. Sim, H.-Q. Yin, H.-S. Choi, Y.-J. Choi, H. C. Kwak, S.-K. Kim, B.-H. Lee, L-Serine Supplementation Attenuates Alcoholic Fatty Liver by Enhancing Homocysteine Metabolism in Mice and Rats. The Journal of Nutrition 145, 260–267 (2014).

40. K. Salic, E. Gart, F. Seidel, L. Verschuren, M. Caspers, W. van Duyvenvoorde, K. E. Wong, J. Keijer, I. Bobeldijk-Pastorova, P. Y. Wielinga, R. Kleemann, Combined Treatment with L-Carnitine and Nicotinamide Riboside Improves Hepatic Metabolism and Attenuates Obesity and Liver Steatosis. Int J Mol Sci 20, (2019).

41. T. M. Hagen, R. T. Ingersoll, C. M. Wehr, J. Lykkesfeldt, V. Vinarsky, J. C. Bartholomew, M. H. Song, B. N. Ames, Acetyl-L-carnitine fed to old rats partially restores mitochondrial function and ambulatory activity. Proc Natl Acad Sci U S A 95, 9562–9566 (1998).

42. S. A. J. Trammell, B. J. Weidemann, A. Chadda, M. S. Yorek, A. Holmes, L. J. Coppey, A. Obrosov, R. H. Kardon, M. A. Yorek, C. Brenner, Nicotinamide Riboside Opposes Type 2 Diabetes and Neuropathy in Mice. Scientific Reports 6, 26933 (2016).

43. P. Bieganowski, C. Brenner, Discoveries of nicotinamide riboside as a nutrient and conserved NRK genes establish a Preiss-Handler independent route to NAD+ in fungi and humans. Cell 117, 495–502 (2004).

44. C. D. Heer, D. J. Sanderson, Y. M. O. Alhammad, M. S. Schmidt, S. A. J. Trammell, S. Perlman, M. S. Cohen, A. R. Fehr, C. Brenner, Coronavirus and PARP expression dysregulate the NAD Metabolome: a potentially actionable component of innate immunity. bioRxiv, 2020.2004.2017.047480 (2020).

45. S. E. Airhart, L. M. Shireman, L. J. Risler, G. D. Anderson, G.A. Nagana Gowda, D. Raftery, R. Tian, D. D. Shen, K.D. O’Brien, An open-label, non-randomized study of the pharmacokinetics of the nutritional supplement nicotinamide riboside (NR) and its effects on blood NAD+ levels in healthy volunteers. PLoS One 12, e0186459 (2017).

46. J. Saleh, C. Peyssonnaux, K. K. Singh, M. Edeas, Mitochondria and microbiota dysfunction in COVID-19 pathogenesis. Mitochondrion 54, 1–7 (2020).

47. J. B. Moore, C. H. June, Cytokine release syndrome in severe COVID-19. Science 368, 473–474 (2020).

48. A. Sahebnasagh, F. Saghafi, R. Avan, A. Khoshi, M. Khataminia, M. Safdari, S. Habtemariam, H. R. Ghaleno, S. M. Nabavi, The prophylaxis and treatment potential of supplements for COVID-19. Eur J Pharmacol, 173530–173530 (2020).

