## Appendix for "Combined Metabolic Activators accelerates recovery in mild-to-moderate COVID-19"

### Supplementary Appendix

This appendix has been provided by the authors to give readers additional information about their work.

Supplement to: Combined metabolic activators accelerate recovery in mild-to-moderate COVID-19

### Study protocol

**Indication**	Coronavirus Disease 2019 (COVID-19)

**Study agent**	Combined metabolic activators consisting of serine, L-carnitine tartrate, N-acetylcysteine, and nicotinamide riboside

**Study phase**	Phase II and Phase III

**Methodology**

Phase II: Open label, randomized controlled design

Phase III: Double blinded, randomized controlled design

**Treatment arm:** At home-treatment with standard therapy + combined metabolic activators consisting of serine, L-carnitine tartrate, N-acetylcysteine, and nicotinamide riboside as compared to standard therapy + placebo. Standard therapy includes only hydroxychloroquine in phase II and either hydroxychloroquine or favipiravir in phase III.

**Primary objective**

Studies are planned as Phase II and Phase III clinical drug research to be conducted in patients diagnosed with COVID-19. Patients will be ambulatory and after the diagnosis/confirmation of diagnosis, will be sent home with their treatment.

The primary objective is to assess the clinical efficacy of the combination of metabolic activators administration and standard therapy in COVID-19 patients.

For the primary purpose, the proportion of patients who were recovered during the course of disease until 14 days after the initial diagnosis of COVID-19 disease.

**Secondary objectives**

The secondary aim is to evaluate the safety and tolerability of combined metabolic activators administration and standard therapy combination.

The following secondary safety objectives have been identified both in phase II and phase III studies:

• Number / characteristics of adverse event (AE), Serious Adverse Event (SAE) and treatment discontinuation due to study drug from the beginning of the study to the end of the follow-up period

• Number / features of all changes in hematology parameters evaluated as AE from the beginning of the treatment to the end of the follow-up period.

• Number / characteristics of all changes in selected biochemical parameters evaluated as AE from the beginning of the treatment to the end of the follow-up period.

• The changes in vital signs (systolic and diastolic blood pressures, pulse, respiratory rate, body temperature, pulse oximetry values), baseline values, and the status of treatment and follow-up visits

**Number of subjects**

A total of 100 COVID-19 disease patients in phase II will be enrolled and randomized on a 3:1 basis to the combined metabolic activators administration + standard therapy or placebo + standard therapy in Turkey. Standard therapy includes only hydroxychloroquine treatment.

A total of 300 COVID-19 disease patients in phase III will be enrolled and randomized on a 3:1 basis to the combined metabolic activators administration + standard therapy or placebo + standard therapy in Turkey. Standard therapy includes either hydroxychloroquine or favipiravir treatment.

**Main inclusion criteria**

To be included in the study, patients should meet all the following criteria:

•	Patients of both genders (females and males) over 18 years of age

•	Written informed consent obtained from the subjects prior to any procedures related to the study.

•	Understand all procedures to be applied within the scope of the study protocol

•	Ambulatory patients with symptoms diagnosed with COVID-19 with real time PCR test result positivity in the last 24 hours

•	Patients with stable clinical course and who could be treated on an ambulatory basis

**Main exclusion criteria**

Subjects cannot be included in the study if any of the following criteria is met:

•	Patients who has partial oxygen saturation below 90% or with severe clinical status requiring admission to critical care unit

•	Inability or unwillingness to give written informed consent

•	Physician makes a decision that trial involvement is not in patients' best interest, or any condition that does not allow the protocol to be followed safely.

•	Patients considered as inappropriate for study for any reason

•	Active participation in another clinical study

•	Uncontrolled Type 1 or type 2 diabetes

•	Severe liver disease (e.g. Child Pugh score ≥ C, AST>5 times upper limit)

•	Patients with known severe renal impairment (estimated glomerular filtration rate ≤30 mL/min/1.73 m2) or receiving continuous renal replacement therapy, hemodialysis, peritoneal dialysis

•	Significant cardiovascular co-morbidity (i.e. heart failure)

•	Patients with phenylketonuria (contraindicated for NAC)

•	Known allergy for substances used in the study

•	Alcohol consumption over 192 grams for men and 128 grams for women per week

•	Pregnant or breastfeeding, or positive pregnancy test in a pre-dose examination

•	Receipt of any experimental treatment for COVID-19 within the 30 days prior to the time of the screening evaluation.

**Dosage and duration of therapy**

Phase II study was planned as a parallel group, randomized and open label study. The study subjects in phase II will be randomized on a 3:1 basis to the combined metabolic activators administration+ standard therapy or placebo + standard therapy in Turkey. Standard therapy includes only hydroxychloroquine treatment. Standard therapy + placebo group will include 25 volunteers; the standard therapy + combined metabolic activators administration group will consist of 75 volunteers.

Phase III study was planned as a parallel group, randomized and double-blinded study. The study subjects in phase III will be randomized on a 3:1 basis to the combined metabolic activators administration+ standard therapy or placebo + standard therapy in Turkey. Standard therapy includes one of the hydroxychloroquine or favipiravir treatment. Standard therapy + placebo group will include 75 volunteers; the standard therapy + combined metabolic activators administration group will consist of 225 volunteers.

In both studies, after the volunteers sign the informed consent forms, the treatment will be administered as the first by the responsible investigator. All other treatments will be administered by the patient at home. Hydroxychloroquine treatment will be administered at an initial dose of 800 mg/day (2x400 mg oral) followed by 400 mg/day (2x200 mg oral) for a total of 5 days. Favipiravir treatment will be administered at a dose of 2x400 mg (oral).

The standard therapy + combined metabolic activators administration group will be applied the same dosage and duration of the standard therapy. Additionally, activators administration mixture will be given for two weeks in doses indicated below (was given orally, twice/day; one dose in the morning, one dose after dinner):

**Drug	Dose**

L-Carnitine tartrate	3.73 g/dose

N-Acetylcysteine	2.55 g/dose

Nicotinamide riboside	1 g/dose

Serine	12.35 g/dose

The total treatment period for the standard therapy will be 5 days and metabolic activators administration will be for 14 days. On the 14th day, patients will perform a last visit which will be the follow-up visit.

**Investigational Products**	Investigational products will be produced as the test product and will be packed as individual dosages in identical HDPE plastic bottles of 60 mL with a screw cap. Test product will be in the form of powder and will be dissolved in 200 ml preferably cold water before use. Placebo treatment will include lactose only in identical HDPE plastic bottles of 60 mL with a screw cap.

**Study design**

The study comprises two visits:

Visit 1 (Screening; Day 0):

The subjects will get the informed consent form and oral information on the study protocol. Blood samples and will be taken to check out the eligibility to the study.

After clinical and physical examination blood samples will be obtained for routine analysis outlined in the study protocol. Chest tomography (CT) and virus load assessment by PCR will be performed. An ECG will also be performed. Plasma samples will be taken for clinical chemistry analysis.

Eligible study subjects will be invited to the clinic for randomization to the active treatment and standard therapy groups. They will be asked to take the first dose and be observed for development of side effects. Patients who can tolerate the study agents will start to take combined metabolic activators (i.e., 2 dosage daily just after breakfast and dinner) for two weeks.

After initial dosing at the hospital patients will be instructed to stay at home during their treatment and will be asked to attend to the investigational site if symptoms are worsened or diminished during the 14-day period.

Visit 2 (Week 2, Day 14):

Clinical and physical examination, adverse events recording, virus load assessment by PCR will be performed. Chest tomography will be performed. Plasma samples will be taken for clinical chemistry analysis.

After the visit 2, subjects will stop taking their study drugs.

**Study duration**

The active treatment duration will be 2 weeks for each subject and the total study duration is estimated as 6 months.

**Efficacy evaluation**

The primary efficacy evaluation will be on the percentage (proportion) of patients recovered during the treatment period for both arms in COVID-19 patients.

The primary objective is to assess the clinical efficacy of the combined metabolic activators administration and standard therapy in COVID-19 patients based on the proportion of recovery.

**Safety evaluation**

Safety will be assessed by monitoring of adverse events, physical examination, vital signs measurements and clinical laboratory tests.

|  | **Visit** | |
| --- | --- | --- |
| **Visits** | **Visit 1** | **Visit 2** |
| **Days** | **Day 0** | **Day 14** |
| Informed consent | X |  |
| Demographic data | X |  |
| Surgical & medical history | X | X |
| Physical examination^1^ and diagnosis | X | X |
| Inclusion/Exclusion criteria | X |  |
| **Laboratory tests** |  |  |
| Blood sample collection^2^ | X | X |
| Chest tomography | X | X |
| ECG | X |  |
| Virus load assessment by PCR | X | X |
| **Efficacy and safety evaluation** |  |  |
| Laboratory safety parameters ^3^ | X | X |
| **Study drug administration** |  |  |
| Randomization ^4^ | X |  |
| Drug administration^5 ,6^ | X |  |
| Monitoring of compliance | X | X |
| Prior & concomitant medication | X | X |
| AE and SAE ^7^ | X | X |

1. Physical examination will include body weight, height, body mass index, vital signs measurements.

2. At both visits, blood samples will be collected for laboratory tests.

3. Laboratory safety parameters will include complete blood count, alanine aminotransferase, aspartate aminotransferase, creatinine, C-reactive protein, triglycerides, cholesterol, glucose, LDH, ferritin, D-dimer.

4. Eligible study subjects at Visit 1 will be offered to be enrolled in the study and to take the first dose at the hospital.

5. Two doses taken just after breakfast and dinner. Patients will be treated on an ambulatory basis. If patients’ symptoms worsen, patient will be instructed to attend the hospital for an examination.

6. The study participants will be observed for the development of any allergic reactions or intolerance after taking the first dose at the hospital.

7. Adverse events (AE) and serious adverse events (SEA) will be monitored continuously and all AEs that occur at any time during the study will be reported in Case Report Forms.

### Table S1. The use of N-acetyl-L-cysteine, Nicotinamide Riboside (Vitamin B3, Niacin) and L-carnitine used in previous human trials associated with viral diseases including COVID-19.

| NCT Number | Title | Status | Interventions | Age  (Years) | Phase | Enrolment | Study Designs | Start Date | Completion Date |
| --- | --- | --- | --- | --- | --- | --- | --- | --- | --- |
| N-acetylcysteine | | | | | | | | | |
| NCT04279197 | Treatment of Pulmonary Fibrosis Due to 2019-nCoV Pneumonia With Fuzheng Huayu | Recruiting | N-acetylcysteine+ Fuzheng Huayu Tablet  N-acetylcysteine | 18 to 65 | Phase 2 | 136 | Randomized Double blinded treatment | Feb-20 | Dec-22 |
| NCT04370288 | Clinical Application of Methylene Blue for Treatment of Covid-19 Patients | Recruiting | N-acetylcysteine Vitamin C  Methylene blue | 18 to 90 | Phase 1 | 20 | Randomized  Open Label  Treatment | Apr-20 | Sep-20 |
| NCT01962961 | N-acetylcysteine to Reduce Oxidative Stress and Improve Endothelial Function in HIV-infected Older Adults | Completed | N-acetylcysteine | 50 and older | Phase 1\|2 | 24 | Randomized Quadruple blinded Treatment | Oct-13 | Oct-15 |
| NCT03900988 | Intravenous N-acetylcysteine and Oseltamivir Versus Oseltamivir in Adults Hospitalized With Influenza and Pneumonia | Not yet recruiting | N-acetyl cysteine | 18 and older | Phase 4 | 160 | Randomized  Triple blinded Treatment | Apr-20 | Jul-22 |
| NCT03281226 | RIPE vs RIPE Plus N-acetylcysteine in Patients With HIV/TB Co-infection | Recruiting | RIPE (2m) and RI (4m)  RIPE+NAC (2m) and RI (4m) | 18 and older | Phase 2 | 50 | Randomized  Open Label  Treatment | Dec-16 | Dec-19 |
| NCT03982537 | Effect of N-Acetyl Cysteine (NAC) on the Oral Microbiome | Not yet recruiting | N-Acetyl-L-Cysteine | 18 and older | Phase 2 | 40 | Randomize  Open Label Supportive Care | May-20 | Oct-23 |
| NCT03967665 | Risk Stratification-directed NAC for Prevention of Poor Hematopoietic Reconstitution | Recruiting | N-acetyl-L-cysteine | 15 to 60 | Phase 3 | 138 | Randomized  Open Label Prevention | Oct-18 | Oct-24 |
| NCT02348775 | Glutathione and Function in HIV Patients | Active, not recruiting | N-acetylcysteine and glycine | 45 to 65 | Phase 1 | 16 | Open Label | Nov-14 | Aug-20 |
| NCT01355198 | Role of HIV on Glutathione Synthesis and Oxidative Stress | Completed | N-acetylcysteine and glycine | 21 to 70 | Phase 1 | 10 | Open Label Treatment | Aug-10 | Sep-11 |
| NCT02930031 | Redox Status and Immune Function | Completed | N-acetylcysteine  Placebo | 18 to 30 | Not Applicable | 10 | Randomized Double blinded | Jan-15 | Mar-16 |
| NCT02080182 | Effect of Acetylcysteine in Pediatric Acute Pyelonephritis. | Completed | Drug: Acetylcysteine  Placebo acetylcysteine | 1 to 16 | Phase 2 | 70 | Randomized Quadruple blinded Treatment | Jan-14 | Apr-16 |
| NCT03460808 | The Combination of Atorvastatin, Acetylcysteine and Danazol as the Treatment of Steroid-resistant/Relapse Immune Thrombocytopenia | Not yet recruiting | Atorvastatin  Acetylcysteine  Danazol | 18 and older | Phase 1\|Phase 2 | 200 | Open Label  Treatment | Mar-18 | Jan-23 |
| NCT04368598 | The Combination of High-dose Dexamethasone and Acetylcysteine as the Treatment of Newly-diagnosed ITP | Recruiting | Dexamethasone\| Acetylcysteine | 18 to 80 | Phase 2 | 44 | Open Label  Treatment | Apr-19 | Dec-20 |
| NCT04545008 | Trial of Famotidine & N-Acetyl Cysteine for Outpatients With COVID-19 | Not yet recruiting | Famotidine\|  N-Acetyl cysteine | 18 and older | Phase 1 | 42 | Randomized  Open Label Treatment | Sep-20 | Aug-21 |
| NCT02688361 | A Bioequivalence Study of an Acetylcysteine 2% Oral Solution Versus a Reference Fluimucil 2% Oral Solution | Completed | Fluimucil 2% solution Acetylcysteine 2% solution | 18 to 45 | Phase 1 | 46 | Randomized  Open Label | Feb-16 | Apr-16 |
| NCT03069300 | N-ACetylcysteine to Reduce Infection and Mortality for Alcoholic Hepatitis | Recruiting | N-acetyl cysteine | 18 and older | Phase 3 | 42 | Randomized  Open Label Treatment | Oct-15 | Jun-25 |
| NCT04455243 | Inflammatory Regulation Effect of NAC on COVID-19 Treatment | Not yet recruiting | N-Acetyl cysteine Placebo | 18 and older | Phase 3 | 1180 | Randomized  Quadruple blinded Treatment | Aug-20 | Aug-21 |
| NCT03236220 | Effect of NAC on the Hematopoietic Reconstitution After Haploidentical Hematopoietic Stem Cell Transplantation | Completed | Drug: N-acetyl-L-cysteine | 15 to 60 | Phase 2 | 35 | Open Label  Treatment | Aug-17 | Dec-18 |
| NCT04419025 | Efficacy of N-Acetylcysteine (NAC) in Preventing COVID-19 From Progressing to Severe Disease | Not yet recruiting | N-acetylcysteine | 18 and older | Phase 4 | 200 | Randomized  Open Label Treatment | Aug-20 | May-21 |
| NCT03197103 | The Impact of N-Acetylcysteine on Volumetric Retention of Autologous Fat Graft for Breast Asymmetry Correction | Unknown status | N-Acetylcysteine | 18 to 40 | Phase 4 | 15 | Randomized  Quadruple blinded Treatment | Jul-17 | May-18 |
| NCT00650091 | Evaluating the Effectiveness of Prednisone, Azathioprine, and N-acetylcysteine in Patients With IPF | Completed | N-acetylcysteine Placebo | 35 to 85 | Phase 3 | 264 | Randomized  Quadruple blinded Treatment | Oct-09 | Jan-14 |
| NCT02249546 | Efficacy of Acetylcysteine-containing Triple Therapy in the First Line of Helicobacter Pylori Infection | Unknown status | N-acetylcysteine + PPI-amoxicillin-clarithromycin  PPI-amoxicillin-clarithromycin | 20 and older | Phase 4 | 654 | Randomized  Open Label Treatment | Sep-14 | Oct-16 |
| NCT00493610 | Mucomyst for Hepatitis C | Suspended | N-acetylcysteine | 18 to 80 | Not Applicable | 5 | Non-Randomized  Open Label Treatment | Nov-06 | Jun-08 |
| NCT01138956 | Immune Response of Visceral Leishmaniasis PatientsTreated With Antimonial Plus N-Acetylcysteine | Unknown status | N-acetylcysteine Pentavalent antimonial | 2 to 50 | Not Applicable | 40 | Randomized Single blinded | Apr-10 | Dec-11 |
| NCT00962442 | N-Acetylcysteine in Severe Acute Alcoholic Hepatitis | Completed | N-Acetylcysteine  placebo | 18 and older | Phase 3 | 44 | Randomized  Open Label Treatment | Sep-00 | - |
| NCT00775476 | Treatment of Systemic Lupus Erythematosus (SLE) With N-acetylcysteine | Not yet recruiting | N-acetylcysteine\|Drug: Placebo | 18 and older | Phase 2 | 290 | Randomized  Quadruple blinded Treatment | Oct-20 | Jun-26 |
| NCT04374461 | A Study of N-acetylcysteine in Patients With COVID-19 Infection | Recruiting | N-acetylcysteine Peripheral Blood | 18 and older | Phase 2 | 86 | Non-Randomized  Open Label Treatment | May-20 | May-21 |
| NCT04458298 | A Study to Evaluate OP-101 (Dendrimer N-acetyl-cysteine) in Severe Coronavirus Disease 2019 (COVID-19) Patients | Recruiting | OP-101  Placebo | 18 and older | Phase 2 | 24 | Randomized Double blinded Treatment | Jul-20 | Nov-20 |
| NCT00397735 | N-acetylcysteine in Intra-amniotic Infection/Inflammation | Completed | Amniocentesis  N-acetylcysteine or placebo | 18 and older | Phase 1\|Phase 2 | 68 | Randomized  Quadruple blinded Treatment | Oct-06 | Aug-18 |
| L-Carnitine | | | | | | | | | |
| NCT03604016 | Study to Assess Efficacy of Besifovir and L-carnitine in Chronic Hepatitis B Patients With Nonalcoholic Fatty Liver | Not yet recruiting | Besifovir dipivoxil  L-carnitine  Tenofovir Alafenamide | 20 and older | Phase 4 | 76 | Randomized  Open Label  Treatment | Sep-18 | Jul-20 |
| NCT02312414 | Effects of Carnitine on Oxidative Stress to IVIR Administration to CKD Patients:Impact of Haptoglobin Genotype | Unknown status | L-Carnitine | 18 to 80 | Not Applicable | 25 | Randomized  Open Label  Treatment | Oct-14 | - |
| NCT01909557 | Acetyl-L-Carnitine Supplementation During HCV Therapy With Peg IFN-Î±2b Plus Ribavirin: Effect on Work Performance. | Completed | Acetyl-L-carnitine | 18 to 90 | Phase 3 | 62 | Randomized  Double blinded Treatment | Jan-10 | Dec-11 |
| NCT01913964 | Acetyl-L-Carnitine Supplementation During HCV Therapy With Pegylated Interferon-Î±2b Plus Ribavirin | Completed | Acetylcarnitine | 45 to 65 | Phase 4 | - | Randomized  Double blinded Treatment | Oct-97 | Oct-97 |
| NCT00225160 | ALCAR Prophylaxis Study | Unknown status | Acetyl L-carnitine | 18 and older | Phase 2 | 50 | Randomized  Double blinded  Prevention | Nov-03 | - |
| NCT00202228 | Lactate Metabolism Study in HIV Infected Persons | Completed | Cofactor supplementation  (thiamine, riboflavin, L-carnitine) | 18 and older | Phase 4 | 30 | Non-Randomize, Open Label Treatment | Jul-02 | Sep-11 |
| NCT00386971 | Effects of L-Carnitine on Postprandial Clearance of Triglyceride-rich Lipoproteins in HIV Patients on HAART | Completed | L-carnitine  placebo | 18 to 70 | Not Applicable | 13 | Randomized  Double blinded Treatment | Oct-06 | Dec-09 |
| NCT00572429 | Effects of Mixed Exercise Regime and L-Carnitine Supplementation in HIV Patients on HAART | Withdrawn | L-carnitine | 18 to 70 | Not Applicable | 0 | Randomized  Double blinded Treatment | Jul-08 | Dec-10 |
| NCT00050271 | Acetyl-L-Carnitine for the Treatment of NRTI-Associated Peripheral Neuropathy | Completed | Acetyl-L-carnitine | 13 and older | Not Applicable | 27 | Randomized  Open Label  Treatment | Jan-07 | . |
| NCT00079599 | L-Carnitine to Treat Fatigue in AIDS Patients | Completed | L-carnitine  Placebo | 18 and older | Phase 2 | 44 | Randomized  Quadruple blinded Treatment | Nov-02 | Mar-07 |
| Nicotinamide riboside (a form of Vitamin B3, Niacin) | | | | | | | | | |
| NCT00246376 | Diet, Exercise, Niacin, and Fenofibrate to Reduce Heart Disease Risk Factors in Individuals With HIV Lipodystrophy or Dyslipidaemia | Completed | Diet  Exercise  Niacin  Fenofibrate | 18 to 65 | Not Applicable | 221 | Randomized  Triple blinded  Treatment | Jan-04 | Feb-12 |
| NCT00046267 | Niacin for Treatment of Elevated Cholesterol and Triglycerides in HIV-Infected Patients | Completed | Niacin | 18 and older | Not Applicable | 30 | Non-Randomized Open Label Treatment | - | - |
| NCT00152893 | To Determine if Chromium Nicotinate Supplementation Will Improve Insulin Resistance in HIV Patients With Metabolic Abnormalities | Completed | Chromium nicotinate | 18 and older | Phase 2 | 52 | Randomized Quadruple blinded Prevention | Aug-02 | Feb-08 |
| NCT01683656 | ER Niacin/Laropiprant Impact on Cardiovascular Markers and Atheroprogression in HIV-infected Individuals on cART | Terminated | Niacin/Laropiprant | 40 and older | Phase 4 | 4 | Randomized Quadruple blinded Treatment | Aug-12 | Jul-14 |
| NCT02018965 | Niacin on Immune Activation : a Proof-of-concept Study | Completed | Niacin | 18 and older | Phase 2 | 16 | Randomized  Open Label Treatment | Nov-11 | Jun-17 |
| NCT01426438 | Endothelial Function, Lipoproteins, and Inflammation With Low HDL Cholesterol in HIV: ER Niacin Versus Fenofibrate | Completed | Niacin  Aspirin  Fenofibrate | 18 and older | Phase 2 | 99 | Randomized  Open Label Treatment | Nov-11 | Oct-13 |
| NCT00986986 | Study of Niacin on Endothelial Function in HIV-infected Subjects With Low High Density Lipoprotein Cholesterol Levels | Completed | Niacin | 18 and older | Not Applicable | 20 | Randomized  Open Label Treatment | Nov-07 | Apr-10 |
| [NCT04271735](https://clinicaltrials.gov/show/NCT04271735) | Pilot Study to Evaluate the Effect of Nicotinamide Riboside on Immune Activation in Psoriasis | Recruiting | Niacin | 18 and older | Phase 2 | 40 | Randomized  Triple blinded Basic Science | May-20 | Sep-23 |
| NCT02812238 | Study to Evaluate the Effect of Nicotinamide Riboside on Immunity | Completed | Nicotinamide riboside (NR) | 18 and older | Not Applicable | 38 | Randomized  Quadruple blinded  Basic Science | Jun-16 | Aug-18 |
| NCT03962114 | Effects of Vitamin B3 in Patients With Ataxia Telangiectasia | Enrolling by invitation | Vitamin B3 | 2 and older | Phase 2 | 24 | N/A  Open Label  Treatment | Mar-19 | Mar-20 |
| NCT00170404 | TB Nutrition, Immunology and Epidemiology | Completed | Folic Acid Micronutrients: Vitamins B1, B2, B6, Niacin, B12, C, E.  Selenium  Vitamin A | 18 to 65 | Phase 3 | 887 | Randomized  Double blinded Treatment | Jun-00 | Oct-05 |
| NCT04407390 | Effects of Nicotinamide Riboside on the Clinical Outcome of Covid-19 in the Elderly | Recruiting | Nicotinamide riboside  Placebo | 70 and older | Phase 2 | 100 | Randomized  Quadruple blinded  treatment | Jun-20 | May-22 |

**Table S2** Baseline demographics of the study population

| **Characteristics** | **Phase-2 Study** | | | **Phase-3 Study** | | |
| --- | --- | --- | --- | --- | --- | --- |
|  | **CMA group** | **Placebo group** | **Study cohort** | **CMA group** | **Placebo group** | **Study cohort** |
|  | **(n=71)** | **(n=22)** | **(n=93)** | **(n=229)** | **(n=75)** | **(n=304)** |
| **Age,** years | 35.0 (19-66) | 32.5 (20-58) | 35.6 (19-66) | 36.7 (18-63) | 35.2 (18-66) | 36.3 (18-66) |
| **Sex** | | | | | | |
| Male | 31 (44%) | 6 (27%) | 56 (60%) | 136 (59%) | 39 (52%) | 175 (57.6%) |
| Female | 40 (56%) | 16 (73%) | 37 (40%) | 93 (41%) | 36 (48%) | 129 (42.4%) |
| **Underlying health conditions** | | | | | | |
| Body-mass index, kg/m^2^ | 24.9 (16.8-37.8) | 24.7 (20.2-33.9) | 24.8 (16.8–37.8) | 27.0 (16.8-45.6) | 25.8 (17.2-34.4) | 26.7 (16.8–45.6) |
| Smoking | 17 (24%) | 6 (27%) | 23(24.7%) | 56 (24%) | 21 (28%) | 77 (25.3%) |
| Alcohol consumption | 3 (4.2%) | 2 (9.1%) | 5(5.3%) | 20 (8.7%) | 9 (12%) | 29 (9.5%) |
| Hypertension | 2 (2.8%) | 0 | 2 (2.1%) | 25 (10.9%) | 3 (4%) | 28 (9.2%) |
| Diabetes Mellitus | 5 (7.0%) | 0 | 5 (5.3%) | 15 (6.6%) | 4 (5.3%) | 19 (6.2%) |
| **Symptoms and signs** | | | | | | |
| Fever | 13 (18.3%) | 6 (27.2%) | 19 (20.4%) | 68 (29.6%) | 18 (24.0%) | 86 (28.2%) |
| Cough | 18 (25.4%) | 11 (50.0%) | 29 (31.1%) | 102 (44.5%) | 30 (40.0%) | 132 (43.4%) |
| Breathing issues | 4 (5.6%) | 2 (9.1%) | 6 (6.4%) | 20 (8.7%) | 8 (10.6%) | 28 (9.2%) |
| Tiredness | 35 (49.3%) | 13 (59.1%) | 48 (51.6%) | 115 (50.2%) | 38 (50.6%) | 153 (50.3%) |
| Muscle or joint pain | 42 (59.2%) | 14 (63.6%) | 56 (60.2%) | 142 (62.0%) | 43 (57.3%) | 185 (60.8%) |
| Headache | 33 (46.5%) | 12 (54.5%) | 45 (48.3%) | 89 (38.8%) | 27 (36.0%) | 116 (38.1%) |
| Smell or taste disorder | 19 (26.8%) | 6 (27.3%) | 25 (26.8%) | 42 (18.3%) | 21 (28.0%) | 63 (20.7%) |
| Sore throat | 19 (26.8%) | 3 (13.6%) | 22 (23.6%) | 59 (25.7%) | 14 (18.6%) | 73 (24.0%) |
| Nausea or vomiting | 5 (7.0%) | 0 | 5 (5.3%) | 18 (7.9%) | 6 (8.0%) | 24 (7.8%) |
| Diarrhea | 2 (2.8%) | 0 | 2 (2.1%) | 6 (2.6%) | 5 (6.6%) | 11 (3.6%) |

Values are n (%) or median (range).

### Table S3. Summary of Univariate Cox regression analysis of overall treatment duration in Phase 2

|  | Phase 2 study | | | |
| --- | --- | --- | --- | --- |
|  | HR | Lower 95% CI | Upper 95% CI | P value |
| CMA | 2.59 | 1.54 | 4.38 | 3.62E-04 |
| HQ or FP | - | - | - | - |
| Age | 1.01 | 0.99 | 1.03 | 2.02E-01 |
| Gender | 0.94 | 0.61 | 1.44 | 7.77E-01 |
| Other treatment history | 0.87 | 0.52 | 1.45 | 6.01E-01 |
| Cigarette usage | 1.29 | 0.80 | 2.08 | 3.05E-01 |
| Alcohol usage | 1.39 | 0.55 | 3.47 | 4.84E-01 |

Note: HR = hazard risk, CI = confidence interval. CMA: Combined Metabolic Activators; HQ: hydroxychloroquine; FP: Favipiravir

### Table S4. Summary of Univariate Cox regression analysis of overall treatment duration in Phase 3

|  | Phase 3 study | | | |
| --- | --- | --- | --- | --- |
|  | HR | Lower 95% CI | Upper 95% CI | P value |
| CMA | 5.63 | 4.11 | 7.71 | <2e-16 |
| HQ or FP | 0.96 | 0.72 | 1.29 | 0.78 |
| Age | 1.00 | 0.99 | 1.02 | 0.427 |
| Gender | 1.33 | 1.05 | 1.67 | 0.0161 |
| Other treatment history | 1.10 | 0.98 | 1.23 | 0.0979 |
| Cigarette usage | 0.95 | 0.73 | 1.24 | 0.72 |
| Alcohol usage | 1.07 | 0.73 | 1.57 | 0.741 |

Note: HR = hazard risk, CI = confidence interval. CMA: Combined Metabolic Activators; HQ: hydroxychloroquine; FP: Favipiravir

**SUPPLEMENTARY** **FIGURE LEGENDS**

**Figure S1.** The effect of combined metabolic activators on the recovery of the patients is presented on the patient groups received standard therapy with **A)** hydroxychloroquine and **B)** favipiravir separately.

**Figure S2.** The effect of hydroxychloroquine and favipiravir on the recovery of the patients is presented.

**Figure S3.** The level of aspartate aminotransferase (AST) **(A),** C-reactive protein **(B)**, platelets **(C)**, white blood cells **(D)**, neutrophils **(E)** and lymphocytes **(F)**, is presented before and after 14 days treatment in the CMA and placebo groups.

**Figure S4.** The level of the level of hemoglobin **(A)**, cholesterol **(B)** and triglyceride **(C)** is presented before and after 14 days treatment in the CMA and placebo groups.

**Figure S5.** **CMA effect the plasma level of lipids**

Plasma level of lipids that are significantly different on Days 14 vs Day 0 in the CMA and placebo groups. **A)** Venn-diagram representing the number of identified lipids that are significantly different on Days 14 vs Day 0 in the CMA and placebo groups. The intersection represents lipids that were altered in both groups. Statistical significance based on paired Student’s t test. FDR value: < 0.001. Association between the plasma level of significantly different lipids **B)** in both groups (n=71); **C)** only in CMA group (n=102) **D)** and only in placebo group (n=5) on Day 14 vs Day 0. Heatmap shows log2FC based alterations in the lipids and asterisks indicate statistical significance based on paired Student’s t test. FDR value: < 0.001. Log2FC, log2(fold change).

**Figure S6.** **CMA effect the plasma level of other metabolites related to metabolism**

Plasma level of metabolites other than amino acids and lipids as well as that are significantly different on Days 14 vs Day 0 in the CMA and placebo groups. **A)** Venn-diagram representing the number of identified amino acids that are significantly different on Days 14 vs Day 0 in the CMA and placebo groups. The intersection represents amino acids that were altered in both groups. Statistical significance based on paired Student’s t test. FDR value: < 0.001. Association between the plasma level of significantly different metabolites **B)** in both groups (n=27); **C)** only in CMA group (n=59) and **D)** only in placebo group (n=9) on Day 14 vs Day 0. Heatmap shows log2FC based alterations in the metabolites and asterisks indicate statistical significance based on paired Student’s t test. FDR value: < 0.001. Log2FC, log2(fold change).

**SUPPLEMENTARY** **TABLES**

**Table S1** Clinical trials used N-acetyl-L-cysteine (NAC), Nicotinamide Riboside (NR) and L-carnitine in treatment of COVID-19 and other viral diseases.

**Table S2** Baseline demographics of the study population

**Table S3** Summary of univariate Cox regression analysis of overall treatment duration in the phase-2 study.

**Table S4** Summary of univariate Cox regression analysis of overall treatment duration in the phase-3 study.

**SUPPLEMENTARY DATASETS**

**Dataset S1** The characteristics of each patient involved in the Phase-2 and Phase-3 studies.

**Dataset S2** Summary of laboratory and physical variables before and after treatment in the CMA and placebo groups in **A)** phase-2, **B)** phase-3 and **C)** all patients.

**Dataset S3** Untargeted metabolomics data for each patient before and after treatment.

**Dataset S4** Plasma level of metabolites that are significantly different on Days 14 vs Day 0 in the CMA and placebo groups. Only metabolites detected in >50% of samples were analyzed.

**Dataset S5** Plasma level of metabolites that are significantly different between the CMA and placebo groups on Days 0 and Day 14. Only metabolites detected in >50% of samples were analyzed.

**Dataset S6** Association between the plasma level all metabolites with the plasma levels of metabolic activators including serine, carnitine, nicotinamide riboside, and cysteine as well as degradation products of hydroxychloroquine including hydroquinone sulfate and 2-methoxyhydroquinone sulfate (1).

**Dataset S7** The Olink multiplex inflammation panel used to detect the dynamic range of 72 proteins in plasma samples of the 93 patients participated in the study.

**Dataset S8** Plasma level of inflammation related proteins that are significantly different on Days 14 vs Day 0 in the CMA and placebo groups. Only proteins detected in >50% of samples were analyzed.

**Dataset S9** Plasma level of inflammation related proteins that are significantly different between the CMA and placebo groups on Days 0 and Day 14. Only metabolites detected in >50% of samples were analyzed.

**Dataset S10** Association between the plasma level all inflammation related proteins with the plasma levels of metabolic activators including serine, carnitine, nicotinamide riboside, and cysteine.

**Dataset S11** Association between the plasma level of clinical variables including ALT, AST, LDH, Triglycerides (TGs) and hemoglobin with plasma level of all metabolites.

**Dataset S12** Association between the plasma level of clinical variables including ALT, AST, LDH, Triglycerides (TGs) and hemoglobin with plasma level of all inflammation related proteins.

**Dataset S13** Multi-Omics Network Data, including edges and nodes information. The network is presented in the iNetModels (http://inetmodels.com).

**Dataset S14** Association between the plasma level of key inflammation related proteins including MCP-4, IL-18R1, HGF, CXCL10 and CSF-1 with plasma level of all metabolites.
