## Supplementary figures and images for "Combined Metabolic Activators accelerates recovery in mild-to-moderate COVID-19"

### FigureS1

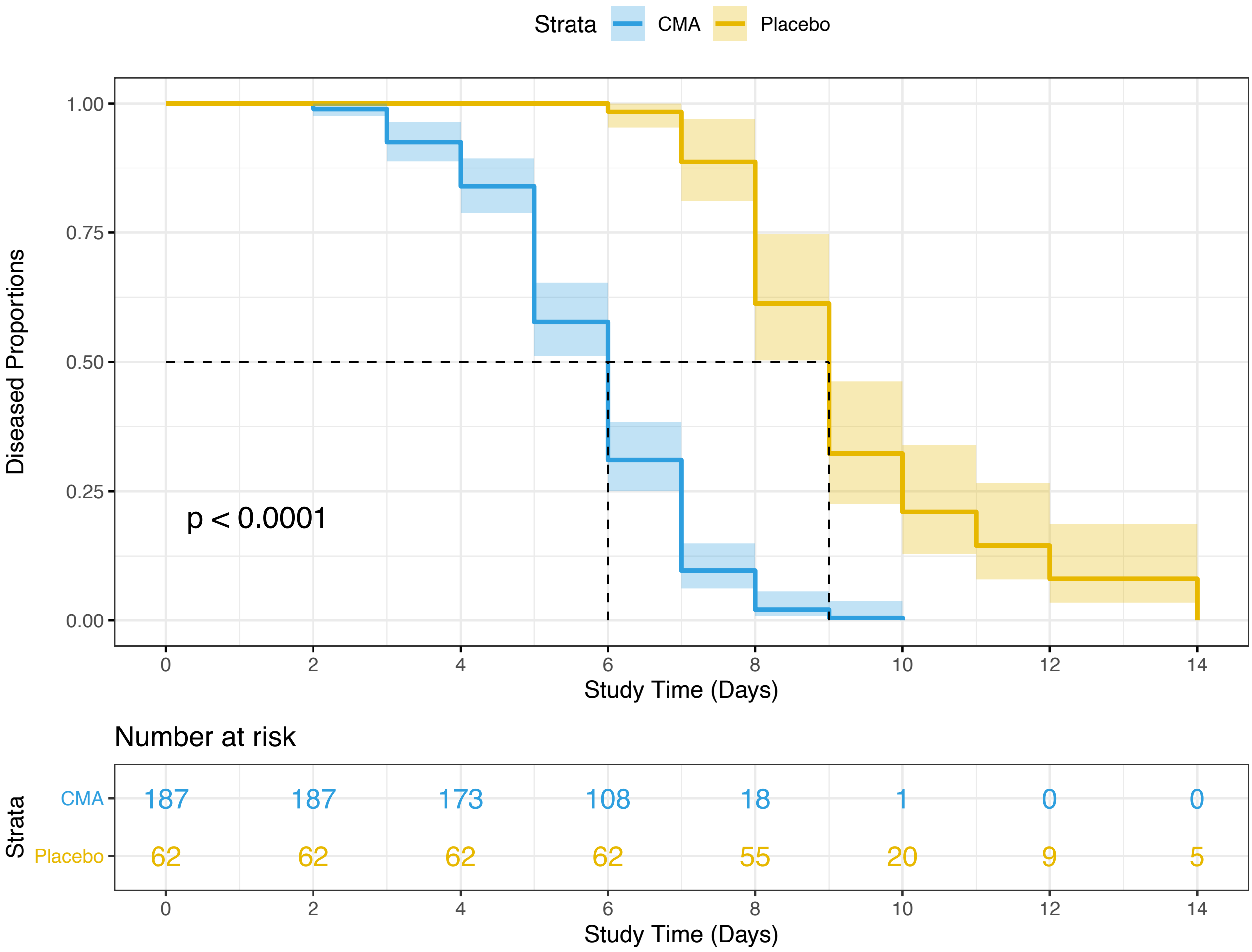

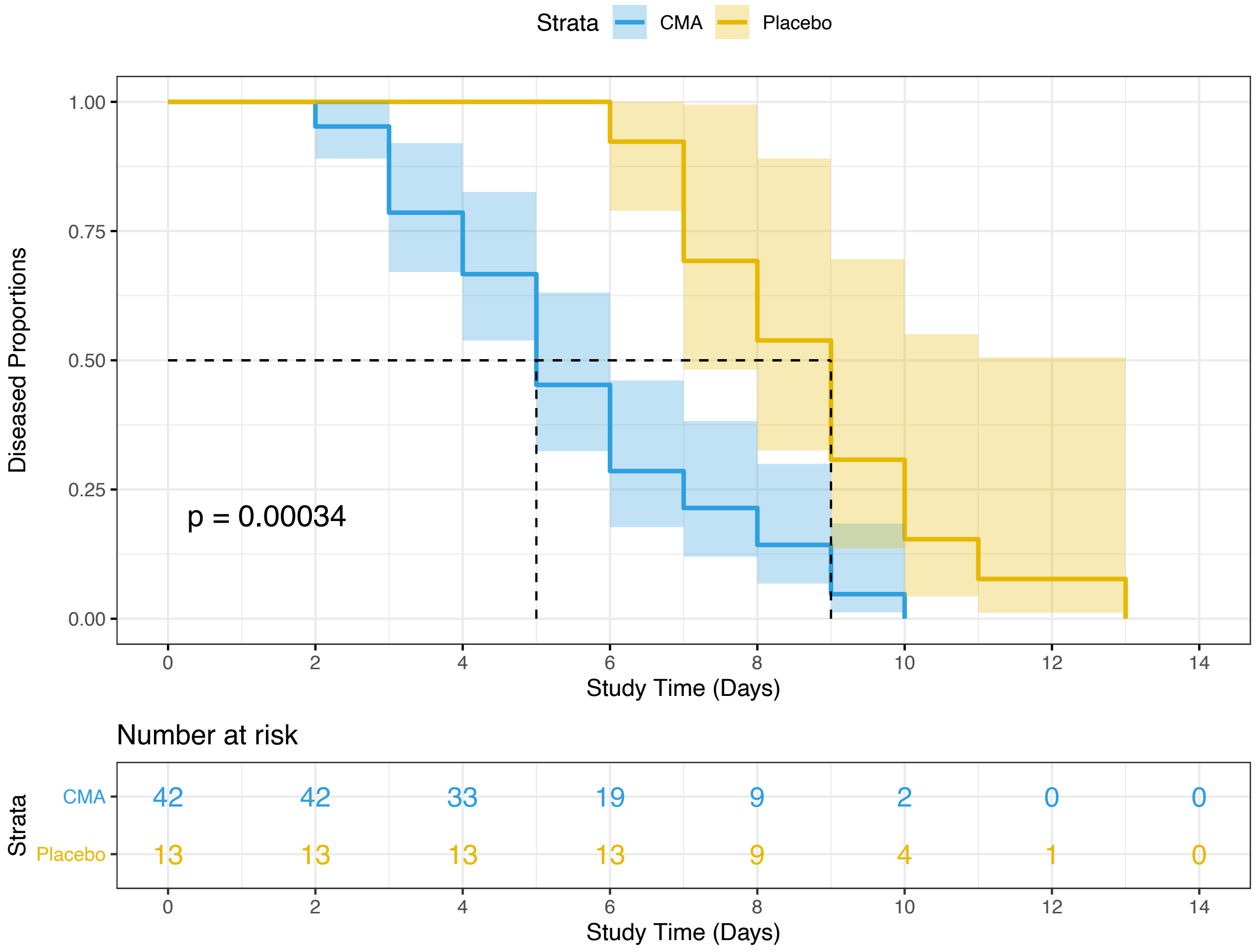

### FigureS2

ALL

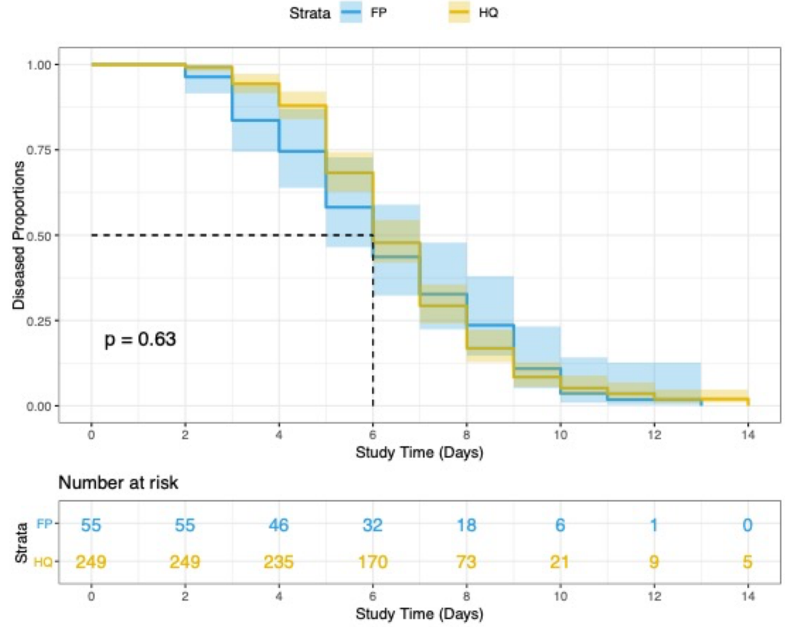


Placebo

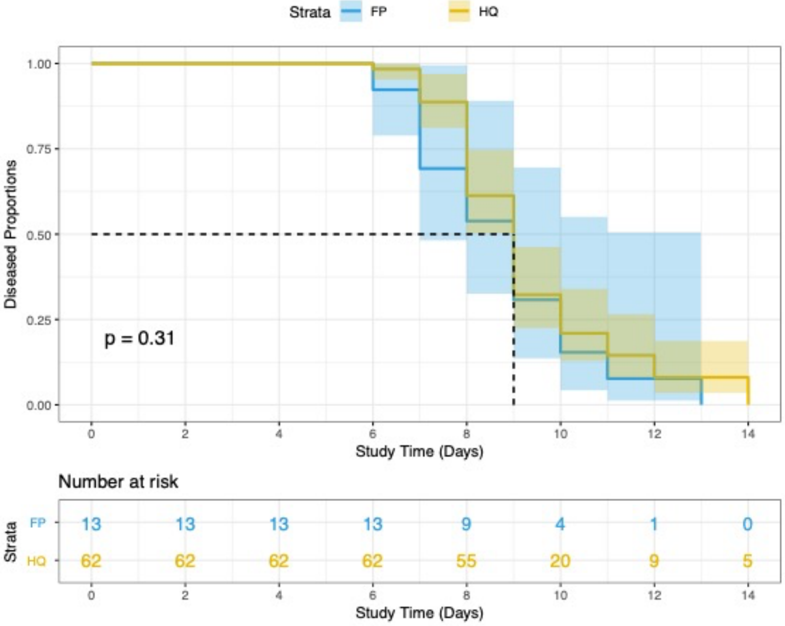


CMA

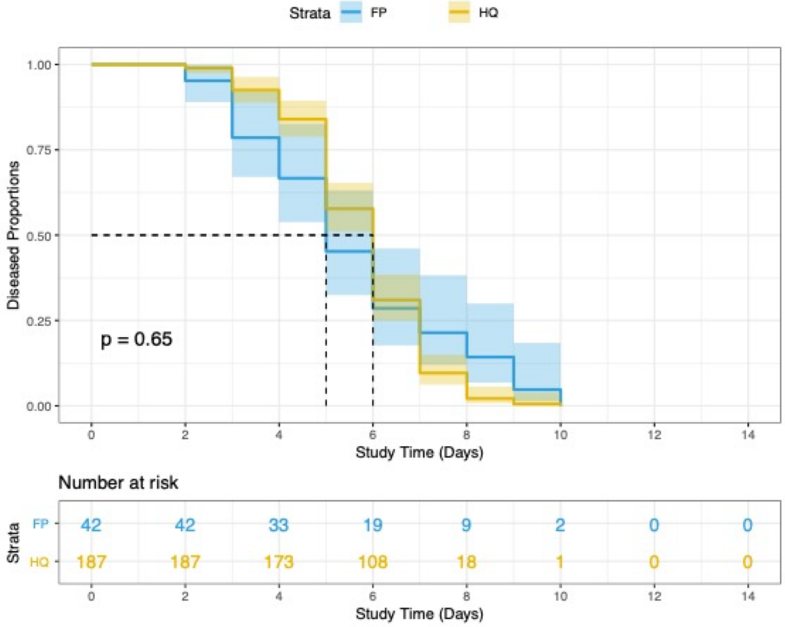

### FigureS3

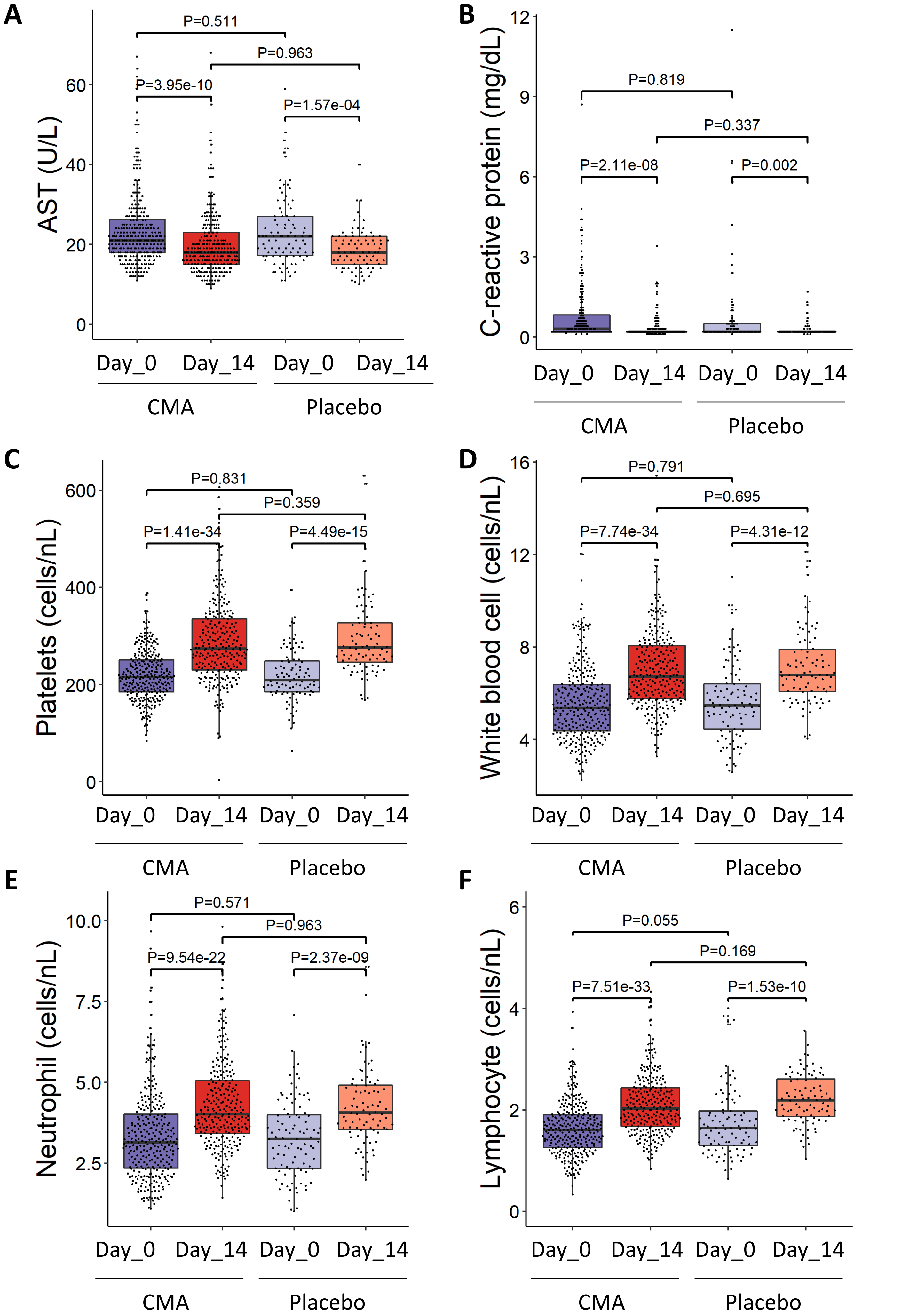

### FigureS4

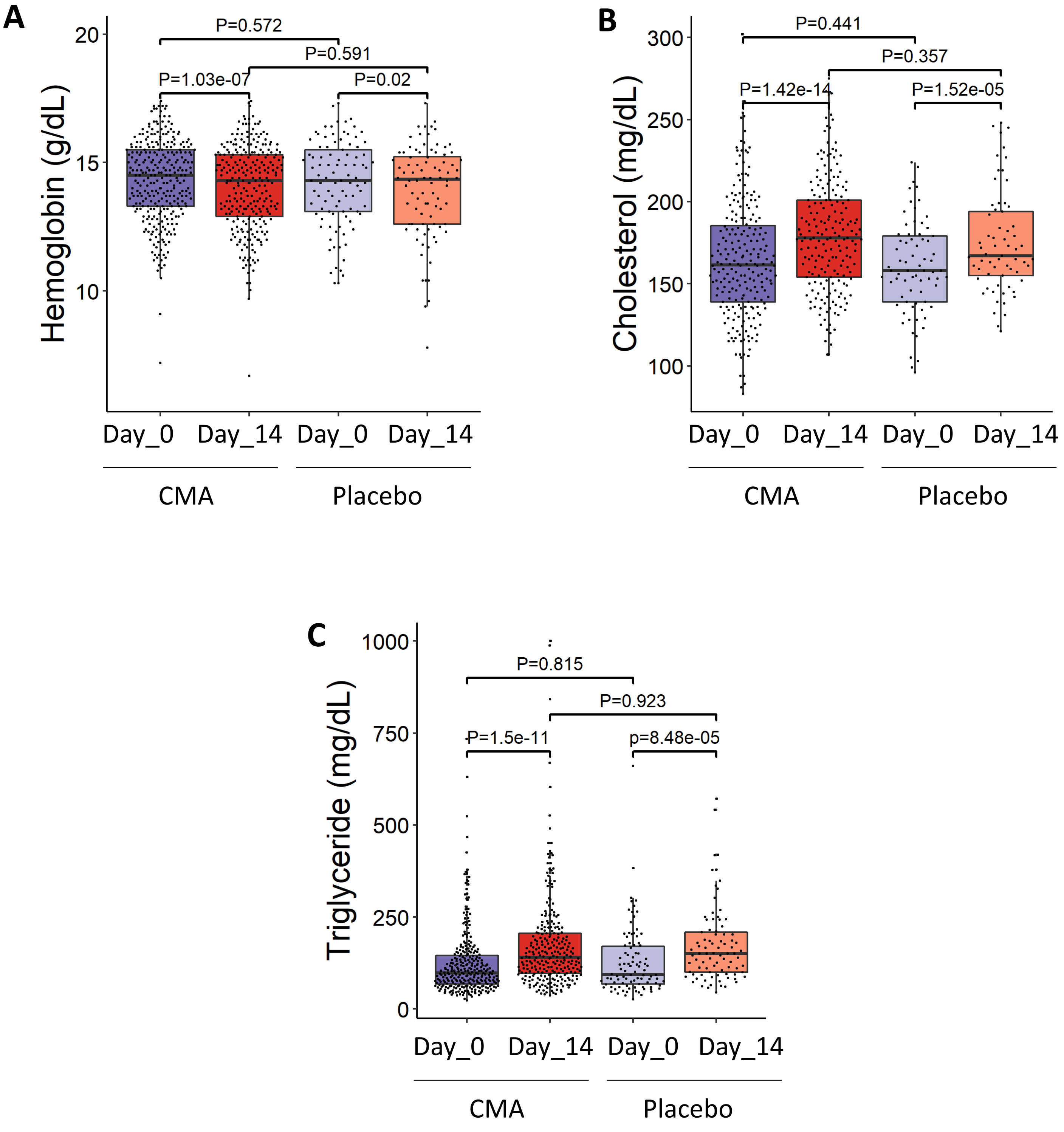

### FigureS5

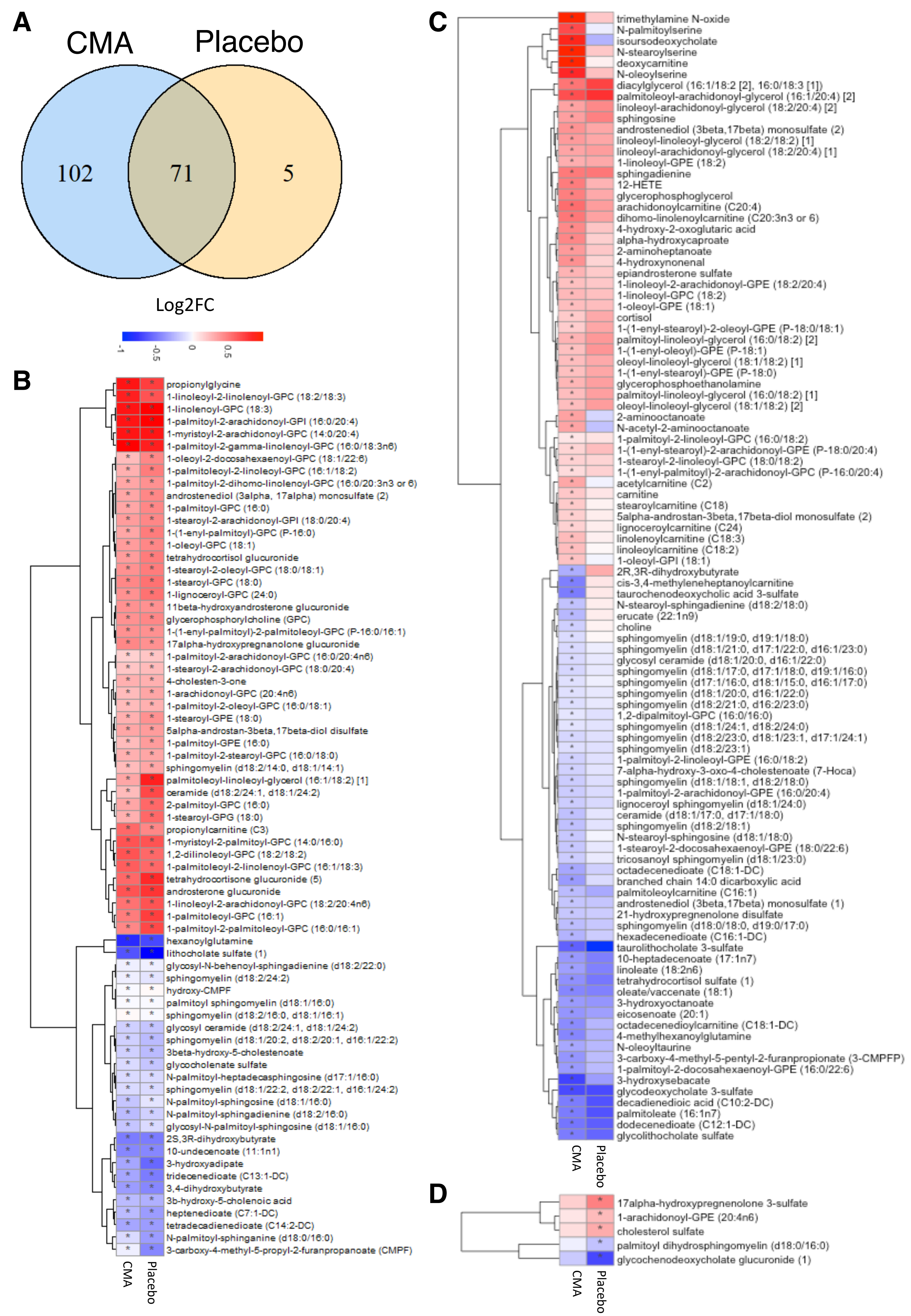

### FigureS6

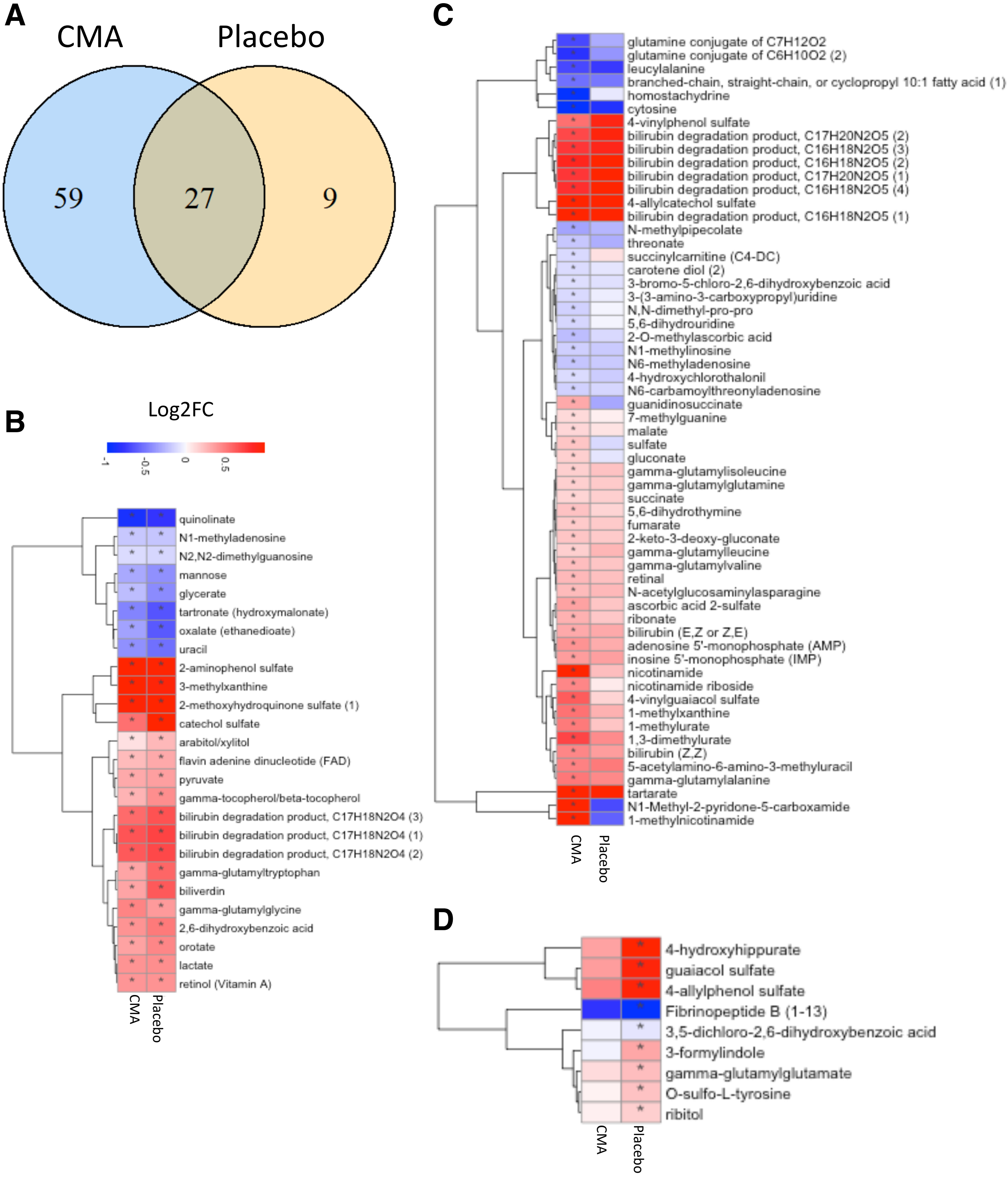
